# Cochlear implants with dexamethasone-eluting electrode arrays reduce foreign body response in a murine model of cochlear implantation and human subjects

**DOI:** 10.1101/2024.10.11.24315311

**Authors:** Muhammad Taifur Rahman, Brian Mostaert, Peter Eckard, Shakila Mahmuda Fatima, Rachel Scheperle, Ibrahim Razu, Bryce Hunger, Rafal T. Olszewski, Shoujun Gu, Cristina Garcia, Nashwaan Ali Khan, Douglas M Bennion, Jacob Oleson, Jonathon R. Kirk, Ya Lang Enke, Robert D. Gay, Robert J. Morell, Keiko Hirose, Michael Hoa, Alexander D. Claussen, Marlan R. Hansen

## Abstract

The inflammatory foreign body response (FBR) following cochlear implantation (CI) can negatively impact CI outcomes, including increased electrode impedances. This study aims to investigate the long-term efficacy of dexamethasone eluting cochlear implant and locally delivered dexamethasone, a potent anti-inflammatory glucocorticoid on the intracochlear FBR and electrical impedance post-implantation in a murine model and human subjects. The left ears of CX3CR1^+/GFP^ Thy1^+/YFP^ (macrophage-neuron dual reporter) mice were implanted with dexamethasone-eluting cochlear implants (Dex-CI) or standard implant (Standard-CI) while the right ear served as unoperated control. Another group of dual reporter mice was implanted with a standard CI electrode array followed by injection of dexamethasone in the middle ear to mimic current clinical practice (Dex-local). Mouse implants were electrically stimulated with serial measurement of electrical impedance. Human subjects were implanted with either standard or Dex-CI followed by serial impedance measurements. Dex-CI reduced electrical impedance in the murine model and human subjects and inflammatory FBR in the murine model for an extended period. Dex-local in the murine model is ineffective for long-term reduction of FBR and electrode impedance. Our data suggest that dexamethasone eluting arrays are more effective than the current clinical practice of locally applied dexamethasone in reducing FBR and electrical impedance.

## 1. Background

A World Health Organization (WHO) 2018 report indicated that hearing loss is the most common sensory abnormality, affecting 466 million (6.1%) of the world population, and predicts that without appropriate intervention, the prevalence of disabling hearing loss will reach 630 million by 2030 and over 900 million by 2050. [1] In addition to the loss of capacity to detect acoustic stimuli, hearing loss negatively affects language and cognitive development, school performance, professional development, and quality of life, and has been identified as the most prevalent modifiable risk factor for dementia. [2–7]

Hair cell loss, the most common cause of sensorineural hearing loss, is caused by aging, acoustic trauma, genetic factors, ototoxins, and inflammation, among other conditions.[8]

Cochlear implants (CI) serve as auditory rehabilitation for sensorineural hearing loss by bypassing the auditory function of the hair cells. Since William House and John Doyle implanted the first single-channel device in 1961 [9], cochlear implant industry has seen tremendous technological advancements in electrode design, programming software, and speech processing strategies. [10]. Cochlear implant electrode arrays are made of platinum/iridium electrodes housed in a silicone elastomer. These materials enable long-term device function and are deemed biocompatible; however, they are not bioinert. [11] and a universal intracochlear foreign body response (FBR) to cochlear implants (CI) has been widely documented. (Claussen, 2022 #966;Claussen, 2019 #836;Nadol, 2008 #896;Noonan, 2020 #900;Seyyedi, 2014 #926) A vigorous FBR following cochlear implantation (CI) is associated with poorer post-implant hearing performance including reduced word recognition score. [12] Further, the extent of the FBR post-CI correlates with electrical impedance across the electrode array. [13–17] [18, 19] Higher electrode impedances require elevated voltages at the electrode-tissue interface potentially leading to reduced dynamic range of stimulation, reduction in clarity, increased energy consumption, and subsequent reduction in battery life. [16]

Although CIs are used in patients with severe to profound hearing loss, preservation of any residual natural acoustic hearing represents an area of intense focus to improve overall performance.[20–22] In particular, preservation of residual acoustic hearing enables combined “acoustic plus electric” (A+E) stimulation, or hybrid stimulation, which markedly enhances CI performance in noisy environments, music appreciation, and sound localization. [23–27] FBR post-CI in animal models and humans has been correlated with post-CI hair cell loss and loss of ‘residual acoustic hearing’. [13, 28–36]

Considering the inflammatory component of FBR, macrophages have been identified as a key contributor following CI in human subjects. [37–39] and animal models. [40–42]. Other than macrophages, T and B lymphocytes have been detected in implanted human cadaveric cochleae.[43] Also, fibrotic tissue response has been detected with *α*-*smooth muscle actin* (*α*-*SMA*) and type 1 collagen [44], markers for contractile fibroblasts and fibrosis, respectively. [45, 46] To mitigate these tissue responses, a non-specific anti-inflammatory compound, dexamethasone has been used extensively clinically, in animal models, and in vitro models in various forms: locally (injection into the cochlea, round window niche, middle ear, dexamethasone eluting implants and rods) and systemically (oral and parenteral) to mitigate FBR post-CI.[47, 48] Sparse clinical data suggest that local dexamethasone in the round window niche, a common clinical practice, reduces electrode impedance. [49] corroborated by animal model data showing reduced FBR[50] and protection of residual hearing.[51, 52] More recently, dexamethasone eluting cochlear implants (Dex-CI) have been developed to provide sustained, intracochlear dexamethasone delivery; early clinical trials have demonstrated that Dex-CI effectively reduce electrode impedance [53] while in animal models, Dex-CI reduces the FBR and electrode impedances and protects hair cells without affecting SGN density. [16, 54–59]

Still, several aspects of the impacts of locally applied dexamethasone and Dex-CI are unknown. First, the diversity of immune cells, and their activation status in implanted cochleae warrant further studies. Second, the long-term pathological changes and impact on the electrical impedance need to be further investigated. Third, the effectiveness of Dex-CI in mitigating the FBR post-CI and reducing electrical impedance in the long term needs to be examined. Fourth, how long-term cochlear implantation affects SGN survival needs to be determined, and how SGN survival is affected by Dex-CI is imperative. Fifth, a comparison between local dexamethasone (a current standard clinical practice) and Dex-CI (an experimental therapy) in reducing FBR post-CI and electrical impedance is pivotal.

## 2. Materials and methods

### 2.1. Experimental animals

All the experimental animal protocols were approved by the University of Iowa Institutional Animal Care and Use Committee, consistent with the Guide for the Care and Use of Laboratory Animals from the Institute for Laboratory Animal Research, National Research Council. Comparable number of both male and female 8–12-week-old CX3CR1^+/GFP^ Thy1^+/YFP^ mice (n=60) on a C57BL/6J/B6 background were used. In these mice macrophages and SGNs express eGFP and eYFP, respectively. [60, 61]

### 2.2. Cochlear implants

#### 2.2.1 Standard and dexamethasone eluting implants (Dex-CI)

For murine implantations, we used standard (comparable to the HL03 electrode array) and dexamethasone-eluting cochlear implants (Dex-CI), provided by Cochlear Limited. While standard implants are previously described in the literature, [40, 62] Dex-CI features a dexamethasone eluting strip embedded in silicone on the intracochlear portion of the electrode array.

#### 2.2.2 Testing dexamethasone content in cochlear implants

Cochlear implants explanted from harvested mice cochlea were analyzed using a Waters Xevo TQ-S cronos triple quadrupole mass spectrometer with an Acquity UPLC H-Class liquid chromatography system. It provides an estimate of the dexamethasone release/eluted in vivo based on a comparison to the results of the t=0 samples. The LC mobile phases were 25 mM Ammonium Acetate with 0.6% Acetic Acid (v/v) in water (Solvent A) and Acetonitrile (Solvent B). A five-minute isocratic LC separation was performed at 40% Solvent B and using a flow rate of 0.4 ml/min. The LC column used was a Waters Acquity BEH C18 (2.1 x 100 mm, 1.7 um) and it was held at 40°C. The injection volume for each sample was 0.5 ul.

The mass spectrometry analysis was performed using positive electrospray ionization and multiple reaction monitoring (MRM).[63] The ESI source parameters used were Capillary voltage 1.5kV, Source temperature 150°C, Desolvation Gas (nitrogen) temperature 400°C; Cone Gas flow 50 l/Hr, and Desolvation Gas flow 800 l/Hr. The three MRM transitions [(M+H)+ to product ions] were used for quantification: 393.13→373.14, 393.13→355.19, and 393.13→147.13. The cone voltage for each MRM transitions were 8V, 26V, and 26V, respectively. The collision energy (eV) used for each transition were 6, 10, and 26, respectively. Waters MassLynx 4.2 software was used for data acquisition and TargetLynx was used for quantitative analysis.

### 2.3. Cochlear implantation in murine model

The timeline for this experiment is shown in Figure 3. Under 1–3% inhalational isoflurane anesthesia, we performed cochlear implantation in the left ear of the mice following the protocol described previously using a round window approach inserting the electrode array to a depth of 2.25 mm. [40, 64, 65] An extended bullostomy was drilled to pack implant lead wire into the tympanic bulla and fixed with dental cement. To limit leakage of perilymph into the middle ear, fascia was packed around the round window. For electrical stimulation and impedance measurements, a transcutaneous connector of the cochlear implant was fed through a subcutaneous pocket from the post-auricular incision to the mid-thoracic spine to be exposed to the external environment.

### 2.4. Single-cell RNA sequencing

#### 2.4.1. Experimental animals

Both male and female 8-12-week-old mice (n = 8; 5 males and 3 females) were implanted in the left ear with standard cochlear implant electrode, with the right ear being unimplanted control.

#### 2.4.2. Single cell suspension

Following CI, mice were euthanized at 33 days post-CI, implanted and contralateral cochlea were microdissected, tissues collected separately into approximately 500 µL DMEM F-12 media and lysed in 3 mL of accutase in a 5 mL tube at 37°C for 30 minutes on a shaker. The media was carefully removed, leaving less than 300 µL of accutase, and replaced with 2 mL of DMEM F-12 containing 5% FBS to stop the lysis. The tissue was triturated for 2 minutes and filtered through a 20 µm filter (pluriSelect Life Science, El Cajon, CA, United States). The filtered cells were then placed in the centrifuge at 300 x g for 3 minutes and resuspended in 90 µL MACS buffer (0.5% BSA in PBS). 10 µL of CD11b microbeads [66-097-142, Miltenyi Biotec, Auburn, Ca, USA] were added to the cell suspension and incubated for 15 minutes at 4°C. Wash step consisting of addition of 1 mL MACS buffer to cell suspension followed by centrifugation step of 350 x g for 5 minutes and subsequent resuspension of cells in 500 µL MACS buffer. Cell suspension was then applied to a prewashed column in a magnetic holder to collect flow-through containing Cd11b negative cells. Column was then washed 3 times with 500 µL MACS buffer with negative flow-through captured each time. The column from the magnet was then removed and placed in a collection tube and then CD11b microbead-bound cells were then eluted with 1 mL MACS buffer. Samples were centrifuged at 300 x g for 3 minutes and concentrated by removing majority of MACS buffer, leaving 90-100 µL at the bottom of the tube with cell pellet. After resuspension by gentle pipetting, cell concentration was measured and adjusted with MACS buffer to 3 × 10^6^ cells/mL. Samples at this concentration were used for 10x cell capture.

#### 2.4.3. 10x Chromium genomics platform

Single-cell captures were performed following the manufacturer’s recommendations on a 10x Genomics Controller device (Pleasanton, CA). The targeted number of captured cells ranged from 3231 to 3572 per run. Library preparation was performed according to the instructions in the 10x Genomics Chromium Single Cell c’ Chip Kit V2. Libraries were sequenced on a Nextseq 500 instrument (Illumina, San Diego, CA) and reads were subsequently processed using 10x Genomics CellRanger analytical pipeline using default settings and 10x Genomics downloadable mm10 genome as previously described. [67]

#### 2.4.4. scRNA-seq data analysis – quality control

Aligned CellRanger count matrix are loaded by Scanpy (v1.9.6) using the function read_10x_mtx. The following cells were filtered in each dataset during the pre-processing steps, respectively:

1. with total read counts below 1 percentile or above 99 percentile;
2. with total genes below 1 percentile or above 99 percentile;
3. with total read counts greater than 10000 or total genes greater than 5000;
4. with mitochondria gene percentage greater than 5%;
5. that are predicted as “doublets” by Scublet with default settings.[68]

Quality metrics are shown in Supplementary Figure S1. Genes by count, total counts, and mitochondrial percentage are shown for cells derived from control (non-implanted) and implanted cochlea (Suppl. Fig. S1A-C). Starting cell counts and ending cell counts after filtering steps are depicted in Supplementary Figure S1D.

#### 2.4.5. scRNA-seq data analysis – annotation

All datasets are normalized using *log1p* normalization. Top 2000 high variable genes are used for Harmony integration of all datasets by Scanpy function *scanpy.external.pp.harmony_integrate* with default settings. The merged dataset is clustered by Leiden algorithm with resolution=1, and annotated based on known marker genes.

#### 2.4.6. scRNA-seq data analysis – differential expression (DE) analysis

DE analysis was performed using DESingle (v3.19) with default settings.

#### 2.4.7. scRNA-seq data analysis – data visualization

All data visualizations were performed using Python packages Matplotlib (v3.7.2) and Seaborn (v0.12.2).

### 2.5. Impedance measurement, neural response telemetry (NRT), and chronic electrical stimulation in murine model

Electrical impedance measurement, nerve response telemetry/NRT (8^th^ nerve electrically evoked compound action potential/ECAP), and programming for chronic electrical stimulation were performed using Custom Sound EP 4.2 according to previously published protocol. Cochlear Ltd., Australia). [40, 64] The Custom Sound programming software uses current level (CL) as its unit ranging between 0 and 255 CL. 0 CL corresponds to 17.5 µAor 0.44 nC/phase and 255 CL corresponds to 1750 µA or 43.75 nC/phase. Impedance and NRT thresholds were measured immediately following surgery, and at least once weekly afterward. During electrical stimulation, electrodes with electrical impedance ≤ 35kOhms were considered within compliance limits and were shorted together using a software patch. Conventional rodent housing was modified with a sliding tether connected to a CI emulator (CIC4 implant emulator, Cochlear Ltd., AUS). Interfacing the receiver coil with a commercial CI sound processor (Cochlear Ltd., AUS), this system was activated. Programmed to 30 CL below the NRT threshold with a dynamic range of 1CL between threshold and comfort levels, electrical stimulation was performed for 5 h per day, 5 days a week, postoperative day 7 through 28 days as described previously. [40]

### 2.6. Immunohistochemistry

Following intraperitoneal injection of ketamine (80mg/kg) and xylazine (10mg/kg), mice were perfused with transcardial ice-cold Phosphate Buffer Solution (PBS) for exsanguination followed by 4% paraformaldehyde (PFA) as fixative. In a rotator, harvested cochleae were fixed overnight with 4% PFA at 4°C, and redundant PFA was removed from samples by incubating overnight in PBS avoiding exposure to light. After decalcification using 0.1M EDTA (pH 7.5) solution, changed every day, for 3-5 days in a rotator, cochleae were rinsed with PBS 3X10 minutes. Cochleae were cryoprotected using serial concentrations (10%-30%) of sucrose solutions. Samples were infused with O.C.T. embedding medium (Tissue-TEK), mounted to the stage of sliding block microtome (American Optical 860) with O.C.T. and dry ice, sectioned parallel to the mid-modiolar plane at 30µm thickness, placed on Fisher Superfrost slides, and stored at −20°C until immunolabeling was performed. For immunolabeling, slides were first warmed to room temperature (∼20-22°C), washed (3 × 5 min each wash) with ‘washing buffer’ containing 0.1% Triton X-100 and 0.3% Tween 20 in TBS and permeabilized and blocked in ‘blocking buffer’ (1% bovine serum albumin (RPI, CAS#9048-46-8) dissolved in washing buffer) for 2 hours. Blocked sections on slides were incubated in primary antibody (Alpha-smooth muscle actin monoclonal antibody, 1A4, eBioscience^TM^, Catalog# 14-9760-82 (1000:1) and MHC Class II (I-A/I-E) Monoclonal Antibody (M5/114.15.2), eBioscience™, Catalog # 14-5321-82 (200:1) in ‘blocking buffer’ overnight (∼16 h) at 4°C. After primary antibody application, sections were washed (3 × 5 min) in ‘washing buffer’, then incubated in blocking buffer containing secondary antibodies Alexa Fluor^TM^ 568, Invitrogen, catalog# **A-11004 (1:400)** and Alexa Fluor^TM^ 750, Novus, catalog# NBP2-68490 **(1:400)** for 2 hours at room temperature. Sections were then washed (3 × 5 min) in ‘washing buffer’. Nuclei were stained with Hoechst 3342 (Sigma) 10 µg/ml in TBS, for 30 min at room temperature, followed by washing with TBS (3X5 min) and cover-slipped with Epredia™ Aqua-Mount Slide Mounting Media (catalog #14-390-5).

### 2.7. Imaging and image analysis

Fluorescently labeled 3 midmodiolar cochlear sections/samples were imaged on a Leica Stellaris 5 confocal system using a 20x (0.70 NA) objective, 0.75x digital zoom, and z-axis-spacing of 1 µm, at an exposure/gain settings to avoid any over/under exposure. Image analysis was performed in IMARIS (Oxford Instruments, UK) image analysis software; cell counts and quantitation of volumetric analyses were done on maximum intensity z-projections of 3D confocal image stacks. Images were coded using a combination of random alphabetical letters and numbers and personnel performing analyses were blinded to the experimental conditions. The outlines of the scala tympani of the basal cochlear turn, Rosenthal’s canal (RC) and the lateral wall in the basal, middle, and apical turns were traced and volume measured with cochlear location defined as previously described. [69] After appropriate thresholding, the number of macrophages (CX3CR1^+/GFP^), neurons (Thy1^+/YFP^), nuclei (Hoechst 3342), and MHCII+ CX3CR1^+/GFP^ macrophages were counted using an automated counting system and density calculated per 10^5^ µm^3^. The fibrotic response was assessed by volumetric quantification of α-SMA in the basal scala tympani in reference to the volume of scala tympani.

### 2.8 Human subjects

Adult cochlear implant candidates were recruited from the University of Iowa Cochlear Implant Clinic and research programs with following exclusion criteria: auditory neuropathy, otosclerosis, large vestibular aqueduct syndrome, Meniere’s or cochlear hydrops, anatomical malformations or involvements of the cochlea/nerve, history of bacterial meningitis, active middle ear infections, currently using ear tubes, unhealed tympanic membrane perforations, known allergy to dexamethasone or corticosteroids. Participants consented to projects approved by the University of Iowa Institutional Review Board (201805740, 202210440, 202307098).

### 2.9 Human cochlear implants

The investigational dexamethasone-eluting array (marked as 632D) is based on the commercially available perimodiolar 632 arrays while the control group was implanted with standard 632 arrays. (Table 1). Drug-eluting silicone wells are distributed along the apical 16 electrodes (numbered 7 thru 22). Due to the limited shelf-life of sterilization of the 632D arrays, participants were not randomized across groups; all 15 632D arrays have been implanted. Recruitment of the control group is ongoing as is longitudinal data collection for both groups.

**Table 1:**
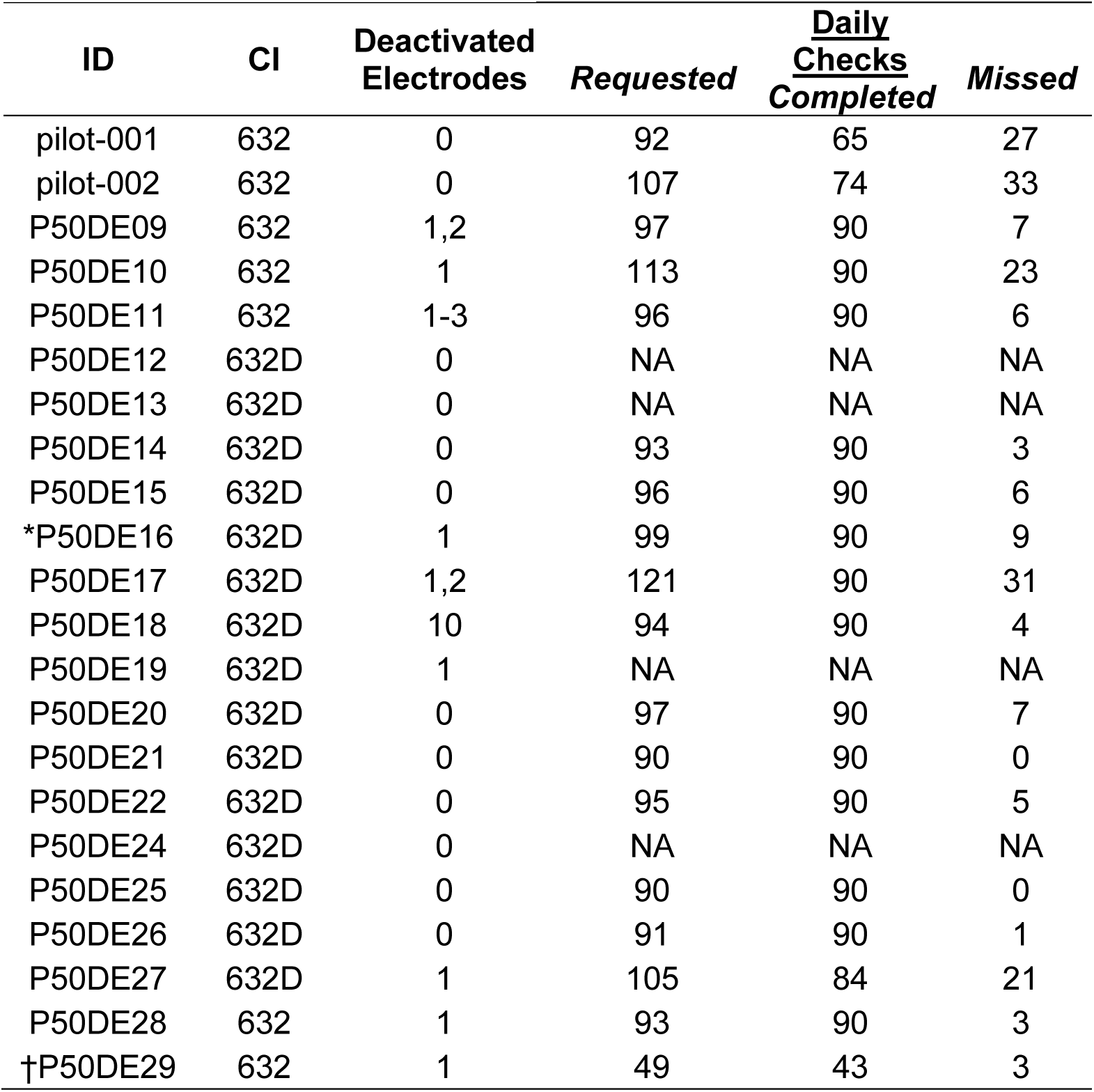
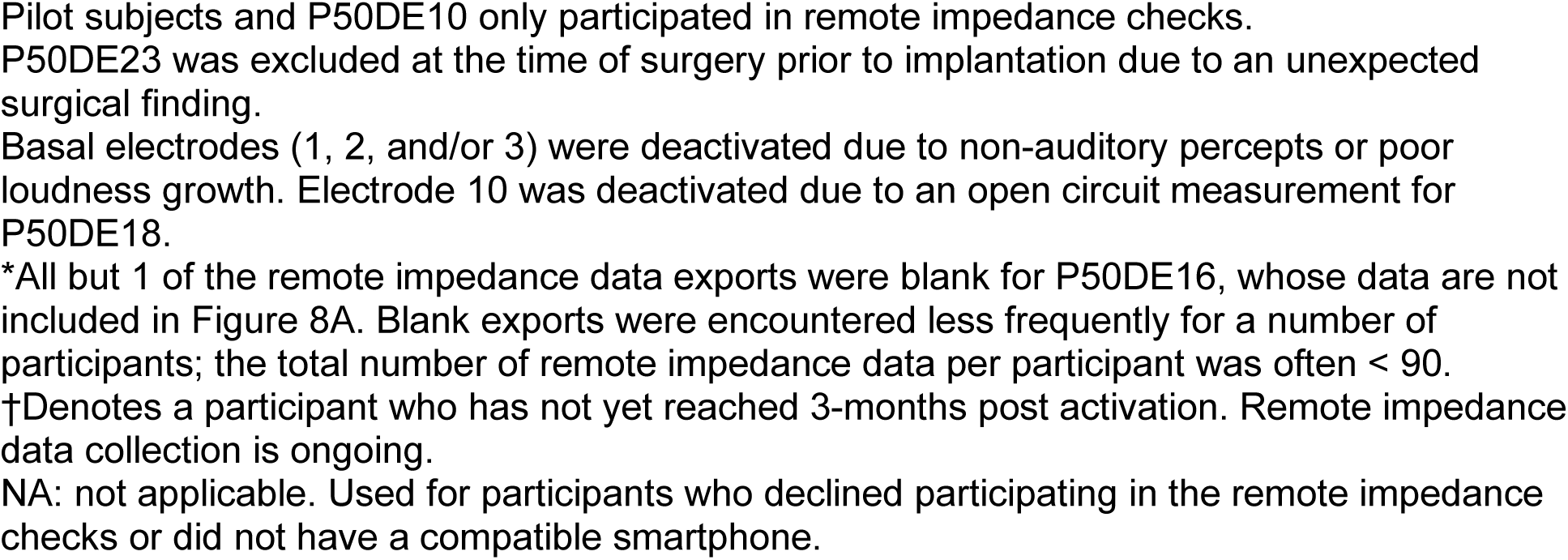
Human subject information.

| ID | CI | Deactivated<br>Electrodes | *Requested* | <u>Daily</u><br><u>Checks</u><br>*Completed* | *Missed* |
| --- | --- | --- | --- | --- | --- |
| pilot-001 | 632 | 0 | 92 | 65 | 27 |
| pilot-002 | 632 | 0 | 107 | 74 | 33 |
| P50DE09 | 632 | 1,2 | 97 | 90 | 7 |
| P50DE10 | 632 | 1 | 113 | 90 | 23 |
| P50DE11 | 632 | 1-3 | 96 | 90 | 6 |
| P50DE12 | 632D | 0 | NA | NA | NA |
| P50DE13 | 632D | 0 | NA | NA | NA |
| P50DE14 | 632D | 0 | 93 | 90 | 3 |
| P50DE15 | 632D | 0 | 96 | 90 | 6 |
| *P50DE16 | 632D | 1 | 99 | 90 | 9 |
| P50DE17 | 632D | 1,2 | 121 | 90 | 31 |
| P50DE18 | 632D | 10 | 94 | 90 | 4 |
| P50DE19 | 632D | 1 | NA | NA | NA |
| P50DE20 | 632D | 0 | 97 | 90 | 7 |
| P50DE21 | 632D | 0 | 90 | 90 | 0 |
| P50DE22 | 632D | 0 | 95 | 90 | 5 |
| P50DE24 | 632D | 0 | NA | NA | NA |
| P50DE25 | 632D | 0 | 90 | 90 | 0 |
| P50DE26 | 632D | 0 | 91 | 90 | 1 |
| P50DE27 | 632D | 1 | 105 | 84 | 21 |
| P50DE28 | 632 | 1 | 93 | 90 | 3 |
| †P50DE29 | 632 | 1 | 49 | 43 | 3 |

Pilot subjects and P50DE10 only participated in remote impedance checks. P50DE23 was excluded at the time of surgery prior to implantation due to an unexpected surgical finding.
Basal electrodes (1, 2, and/or 3) were deactivated due to non-auditory percepts or poor loudness growth. Electrode 10 was deactivated due to an open circuit measurement for P50DE18.
\*All but 1 of the remote impedance data exports were blank for P50DE16, whose data are not included in Figure 8A. Blank exports were encountered less frequently for a number of participants; the total number of remote impedance data per participant was often < 90.
†Denotes a participant who has not yet reached 3-months post activation. Remote impedance data collection is ongoing.
NA: not applicable. Used for participants who declined participating in the remote impedance checks or did not have a compatible smartphone.

### 2.10 Cochlear implantation in human subjects

Subjects underwent unilateral cochlear implantation with either the 632D investigational device or FDA-approved 632 device using standard Instructions for Use by a board-certified neurotologist. Standard facial recess approaches for round window or cochleostomy array insertion were used, depending on surgeon preference. Subjects in both groups received intravenous dexamethasone (10mg) peri-operatively, but no additional corticosteroid drugs were administered in the peri- or immediate post-operative period.

### 2.11 Remote Impedance Measurements in human subjects

Remote impedance measurements were initiated on or shortly after the initial activation appointment following cochlear implantation. Patients/devices were registered in the Cochlear™ online patient portal and paired their hardware to Cochlear™ Nucleus® Smart App. Upon daily scheduled measurement request by a research team member via the app, participants completed daily remote impedance checks. Email reminders were sent when a measurement was not completed, and participants were asked to report technical issues to the team. The target number of daily checks was 90. Adherence varied across participants and some of the daily data were lost due to technical issues. Table 1 provides more details regarding the data available per participant.

### 2.12 Clinic Impedance Measures

Willing participants were seen by the research team at the time of surgery and at each clinical post-operative follow-up programming appointment. Electrode impedance was measured in CustomSound EP via the transimpedance matrix [70] module. Stimulation current was 90-CL with a pulse phase duration of 50 µS. The return path for the stimulus used the ECE1 (rod) extracochlear electrode. The return path for the recording used the ECE2 (case) extracochlear electrode. Only on-electrode recordings using the first phase of the stimulating current pulse are reported. The voltage measured at 6 µS was considered to be essentially instantaneous and used to estimate the real or resistive component (i.e. access resistance). Even though the stimulating current level is constant for the duration of the pulse, recorded voltage increases over time with a shallower slope following the steep onset. This delayed voltage increase is used as the estimate of the reactive component (i.e. polarization impedance). To calculate the polarization impedance, voltages measured at 50 µS were subtracted from voltages measured at 6 µS. [19, 34]

### 2.13. Statistical analyses

Statistical analyses for counts of immune cells, nuclei, neurons, and volume of fibrotic tissue within scala tympani were performed using GraphPad Prism 10.2.2. Specific comparisons that were made are described in respective figure legends. Shapiro-Wilk test was used to determine the normality of data. A two-way ANOVA with Tukey’s multiple comparison was used while the Multiple Mann-Whitney test was applied for data that were not normally distributed. One-way ANOVA with Bonferroni’s multiple comparisons was used for analysis of UPLC data.

A linear mixed model is used to evaluate changes in impedance over time. The fixed effects in the model are group (Standard, Dex CI, Dex Local), days, and an interaction between group and days. The interaction allows the groups to grow at different rates. Since the trend over time appears nonlinear, we model the square root of days. The random effect included in the model is a random intercept for the subject which accounts for repeated observations per mouse. The model was fit using the lme4 package in R. Statistical significance was defined as p<0.05).

## 3. Results

### 3.1. Single cell RNA sequencing data shows diversity of cellular infiltration into cochlea following cochlear implantation

A diverse array of infiltrating immune cells is seen from MACS-based capture and subsequent single cell RNA-sequencing, including monocytes, macrophages, B cells, T cells, neutrophils (Csf3r+, Ly6g+), and red blood cells (Fig. 1A). The distribution of cells derived from implanted cochlea in red and control (contralateral) cochlea in blue can be seen in Figure 1B. The monocyte population can be further clustered into 3 subpopulations (Fig. 1C). Cell counts for each immune cell population detected are shown in Figure 1D. The percent distribution and ratio of immune cells from implanted and control cochlea are shown in Figure 1E and 1F, respectively. Note that few B and T cells and red blood cells (RBCs) were sequenced likely based on the method utilized for MACS-based cell isolation with a focus on assaying monocytes and macrophages amongst the infiltrating immune cell population.

**Figure 1:**
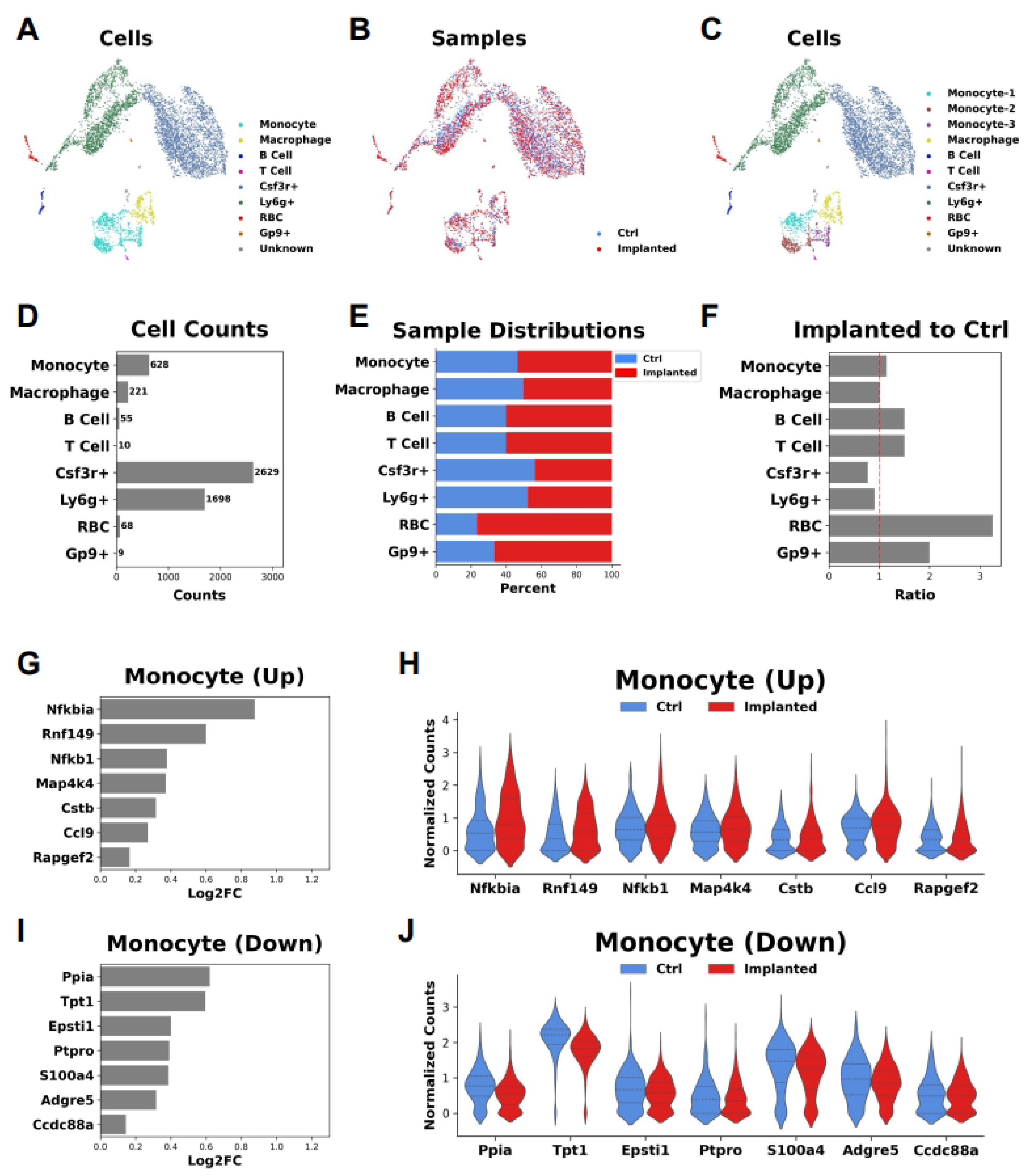
scRNA-Seq reveals a diverse group of immune cells in the cochlear microenvironment with an increase in infiltrating monocytes in the setting of a cochlear implant. A, UMAP plot demonstrates a diverse array of immune cells including monocytes, macrophages, B cells, T cells, and neutrophils where each cell is represented by a dot and transcriptional similar groups of cells cluster together and are color-coded for ease of identification. **B,** UMAP plot depicts the distribution of cells derived from implanted (red dots) and control (non-implanted) cochlea. **C,** UMAP plot demonstrates further subclustering of the monocyte population into 3 subpopulations. **D,** Bar plot depicts immune cell population counts. **E,** Percent distribution of immune cells from implanted (red) and control (blue) cochlea. **F,** Ratio of immune cells from implanted compared to control cochlea depicts increases in monocyte populations. **G,** Differential expression analysis shown in the bar plot demonstrates the top upregulated genes in monocytes derived from implanted cochlea compared to control cochlea. **H,** Violin plots show expression levels (in normalized counts) in these top upregulated genes in both monocytes derived from control (blue) and implanted (red) cochlea. **I,** Similarly, differential expression analysis shown in the bar plot demonstrates the top down-regulated genes in monocytes derived from implanted cochlea compared to control cochlea. **J,** Violin plots demonstrate expression levels (in normalized counts) in these top down-regulated genes in both monocytes derived from control (blue) and implanted (red) cochlea.

Subsequently, differential expression analysis of the transcriptome of monocytes between implanted and control cochlea was performed (Fig 1G-I). Genes involved in suppression of the inflammatory response, including *Nfkbia* and *Nfkb1*, were upregulated in implanted cochlea (Fig. 1G-H). [71–74] Bar plots demonstrate expression fold change for each gene comparing expression in monocytes from implanted cochlea to control cochlea and violin plots demonstrate relative expression of each gene in the monocytes from implanted and control cochlea. In contrast to the above, *Map4k4* expression (Fig. 1G-H), which has been shown to have a proinflammatory effect, is upregulated in monocytes derived from implanted cochlea, suggesting that the regulatory picture may be more complex. [75–78] Further supporting this complex picture is the downregulation of *Tpt1*, *Epsti1*, *S100a4*, and *Adgre1* (Fig. 1I-J). Tumor protein translationally-controlled 1 (*Tpt1*), encodes TCTP (previously known as histamine-releasing factor or HRF) has been shown to promote allergy-associated inflammation.[79]

Epithelial stromal interaction 1 (*Epsti1*) has been shown to be highly expressed in activated macrophages and its deficiency in bone marrow-derived macrophages results in an enhancement of the M2 macrophage phenotype.[80] S100 calcium binding protein A4 (*S100a4*) plays a role in amplifying an inflammatory microenvironment contributing to nuclear factor (NF)-ĸB activation in macrophages in the setting of colon inflammation.[81] Adhesion G-protein-coupled receptor E1 (*Adgre1*) expression is associated with monocyte-to-macrophage transition[82, 83] and immune cell infiltration in head and neck tumors.[84] Overall, these findings support the idea that monocytes may be acting in response to an inflammatory trigger (cochlear implant).

Examining for the possibility of a change in monocyte signature, we performed a deeper examination of the monocyte subcluster transcriptional profiles. Monocyte populations were subclustered separately and 3 subpopulations were identified (Fig. 2A). Differential expression analysis identified top differentially expressed genes in each of the 3 monocyte subpopulations (Fig. 2B-E). Transcriptional changes with respect to differential gene expression seen in monocytes as a group was similar across monocyte subpopulations.

**Figure 2:**
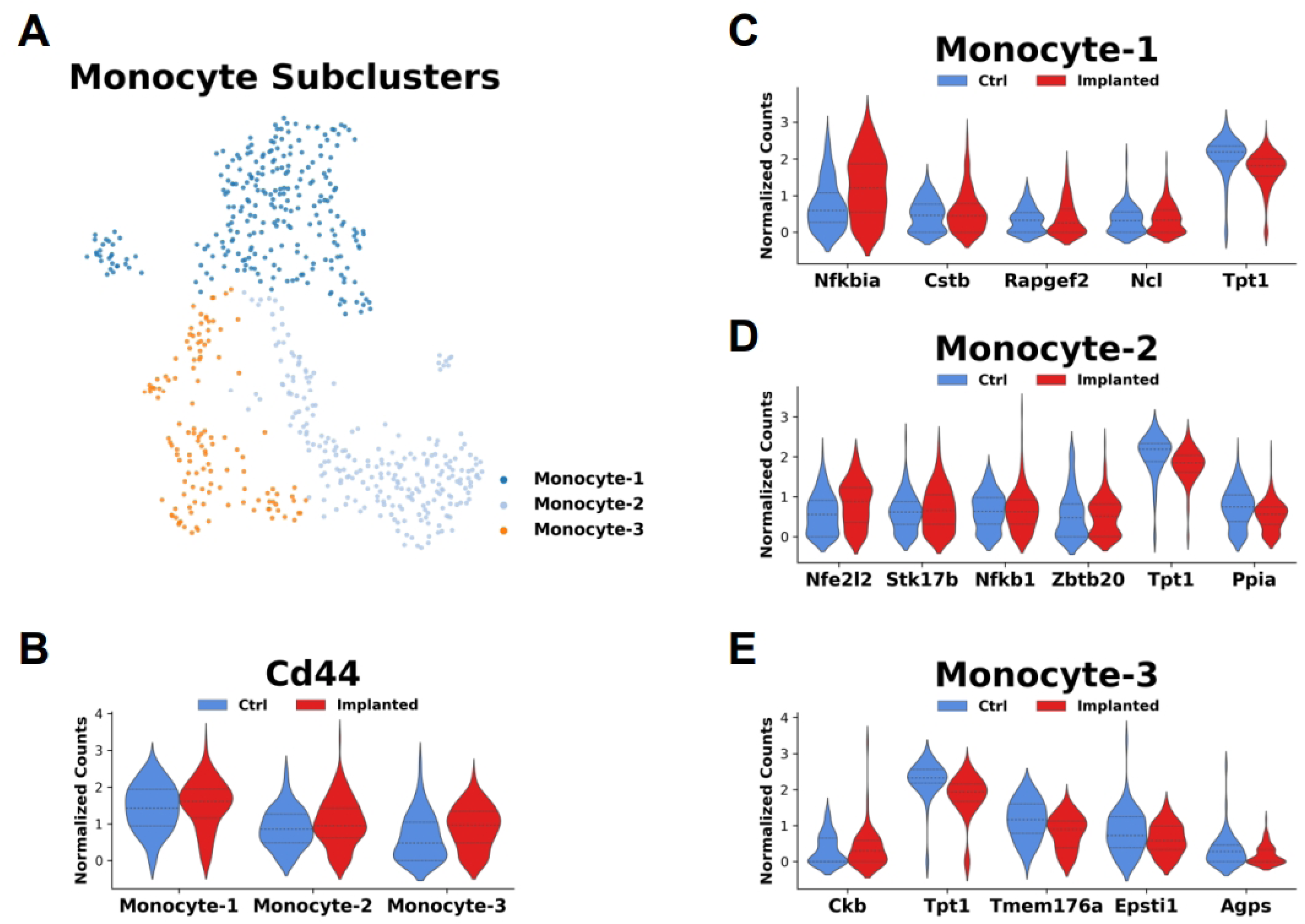
Transcriptional analysis of infiltrating monocytes suggests shift to M1 macrophage phenotype. A, UMAP demonstrates monocyte subpopulations (Monocyte 1-3). **B,** Violin plot demonstrates similar CD44 expression amongst monocyte subpopulations. **C,** Violin plot demonstrates top differentially expressed genes in monocyte-1 subpopulation. **D,** Violin plot demonstrates top differentially expressed genes in monocyte-2 subpopulation. **E,** Violin plot demonstrates top differentially expressed genes in monocyte-3 subpopulation. **F,** Box-and-whisker plots demonstrate relative change in M1 and M2 expression profiles in the monocyte subpopulation between implanted (red box) and control (blue box) with statistically significant difference in the M1 expression profile in monocytes derived from implanted cochlea compared to control (p<.01). **G,** Box-and-whisker plots in the monocyte-1 subpopulation demonstrate similar difference in M1 expression profile between implanted and control cochlea (p<.05) with no difference in M2 expression profile. **H,** Box-and-whisker plots in the monocyte-2 subpopulation demonstrate no difference in M1 or M2 expression profile between implanted and control cochlea. **I,** Box-and-whisker plots in the monocyte-3 subpopulation demonstrate no difference in M1 or M2 expression profile between implanted and control cochlea.

### 3.2. Dexamethasone eluting implants contain dexamethasone for an extended period in the murine model

As shown in Figure 3, once implanted, a significant decrease in the dexamethasone content is observed in Dex-CIs suggesting that dexamethasone is released in-vivo within the implanted cochlea(p=0.013,0.0076,0.0008 at 10, 56 and 112-days post-CI). (Ordinary one-way ANOVA with Bonferroni’s multiple comparison test)

**Figure 3:**
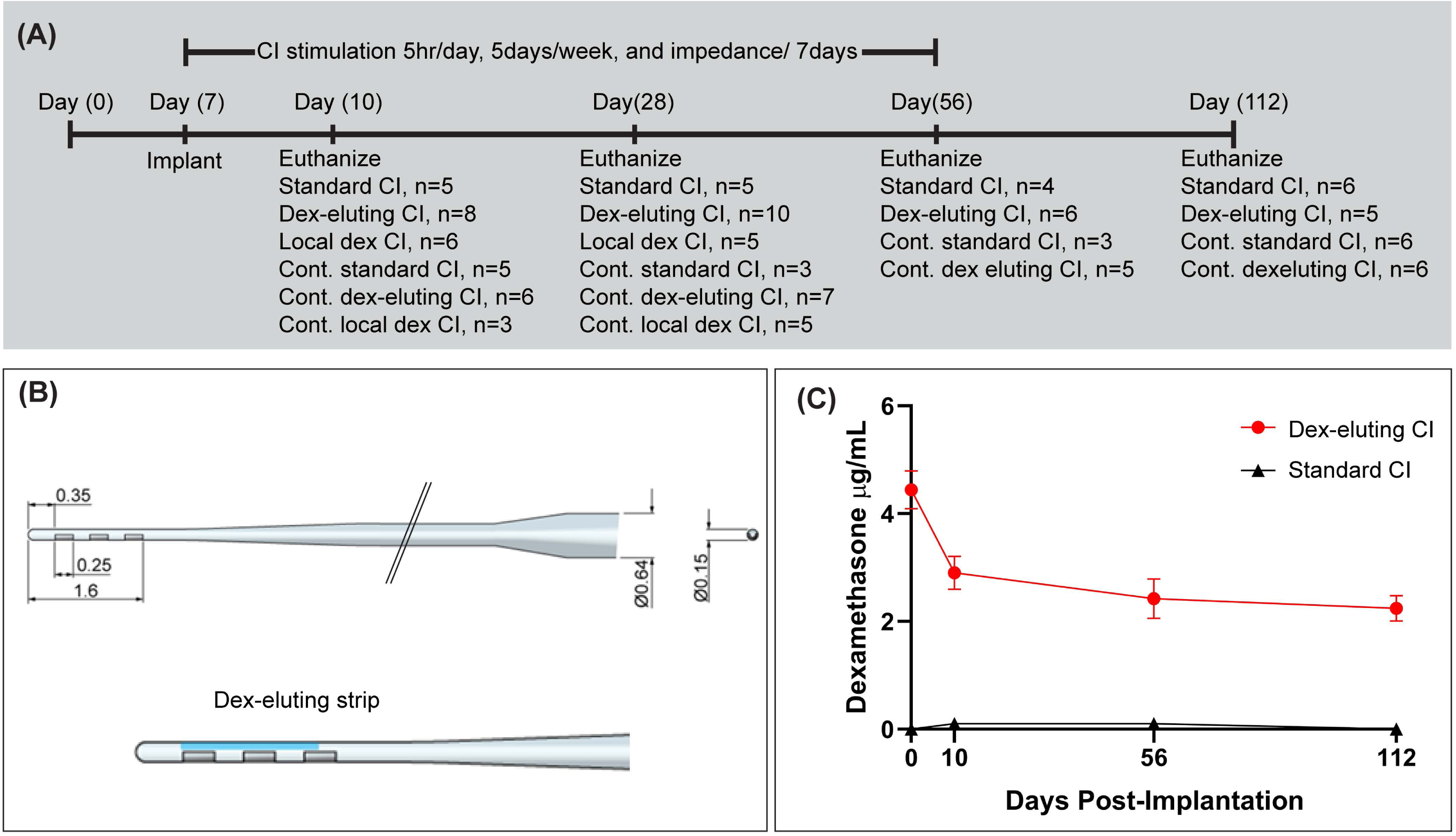
Dexamethasone eluting implant study overview: A. Experimental design for dexamethasone eluting implant study: The number of cochlea (n) used for histopathology has been mentioned. B. Schematic representation of standard cochlear implants: The electrode assembly of the standard cochlear implant consists of a half-banded, three-contact intracochlear array: 2.25 mm in length, 0.15 mm in width at the tip, and 0.64 mm in the widest part (base). The distance between the tip and the apical electrode (E3) is 0.35 mm. Each electrode has a length of 0.25 mm. The distance between the tip of the implant and the basal end of the first electrode (E1) is 1.6 mm. All the parameters shown in the figure are in mm. The intracochlear array tapers to a wider extracochlear helix lead wire insulated with silicone. The intracochlear and two extracochlear electrodes are connected to a transcutaneous 6-pin connector. C. Schematic representation of dexamethasone eluting implants: Structurally, dexamethasone eluting implants are comparable to the standard implants except a dexamethasone-eluting strip (blue) is attached to the apical intracochlear part of the implant. D. Dexamethasone content of cochlear implants explanted from implanted mice: Following euthanasia of the implanted mice, 4 mm of the electrode from the tips of the cochlear implants was collected at 10-, 56-, and 112 days post-CI. Remaining dexamethasone content was measured using UPLC-MS. Error bars indicate SEM. Statistical analysis was performed using Ordinary one-way ANOVA with Bonferroni’s multiple comparison test.

### 3.3. Dexamethasone-eluting cochlear implants reduce the density of CX3CR1+ macrophages in the base of the cochlea in the murine model

Figure 4 shows basal turn from representative mid-modiolar sections across groups and time points. Quantification of CX3CR1-positive cell density in different regions of cochlea across the study period is shown in Figure 5. In the base of the cochlea, standard CI causes the recruitment of CX3CR1-positive cells until 56 days post-CI into the scala tympani (p=0.007, 0.0021, and 0.0001, at 10-, 28- and 56 days post-CI, respectively) and until 28 days in the lateral wall (p=0.0043, 0.0104 at 10- and 28-days post-CI respectively) and spiral ganglion (p=0.0038 and 0.0024 at 10- and 28-days post-CI respectively). Dex-CIs dramatically reduced the CX3CR1+ macrophage density until 56 days post-CI in scala tympani and lateral wall (for both areas, p=0.0001, 0.0001, and 0.0001 at 10-, 28- and 56-days post-CI, respectively). For the spiral ganglion at the base of the cochlea, Dex-CI continued to keep the macrophage density significantly lower than standard CI until 112 days post-CI (p=0.0001, 0.0001, 0.001, and 0.0096 at 10-, 28-, 56- and 112-days post-CI, respectively). All the analyses used a two-way ANOVA with Tukey’s multiple comparison test.

**Figure 4:**
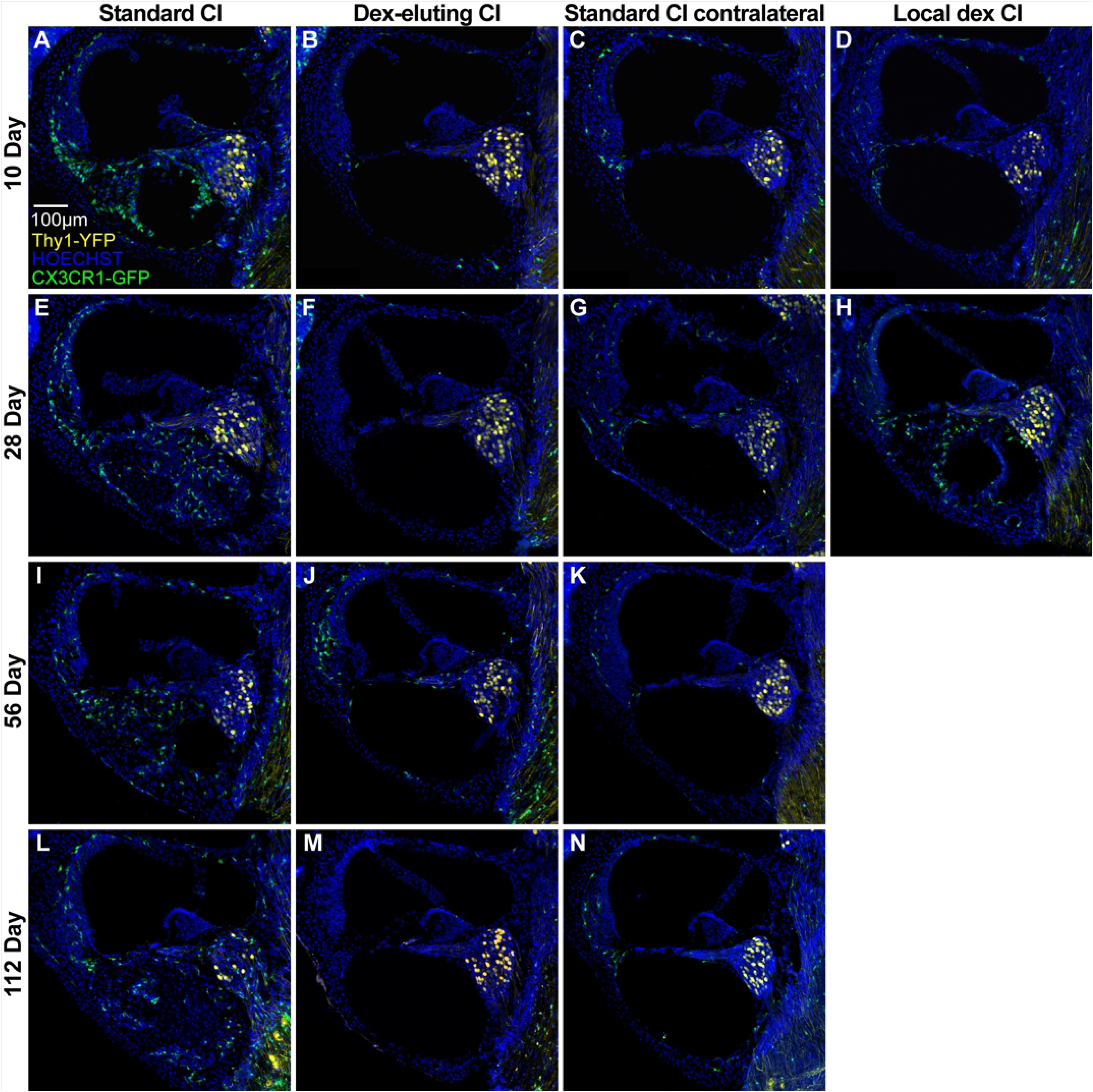
Dexamethasone eluting implant reduces CX3CR1+macrophage and cellular infiltration in basal cochlear turn CX3CR1^+/eGFP^ Thy1^+/eYFP^ mice were implanted with either a Standard cochlear implant (Standard CI) or Dexamethasone eluting Implant (Dex-CI). In a subset of the Standard CI group, Dexamethasone was injected into the round window niche (Dex-local). All implants were electrically stimulated, and mice were euthanized at 10, 28, 56, or 112 days. A-N. Maximum intensity z-projections of 3D confocal image stacks taken from 30-µm thick, midmodiolar sections showing the basal turn of the cochlea. Asterisks show the tract of the CI.

**Figure 5.**
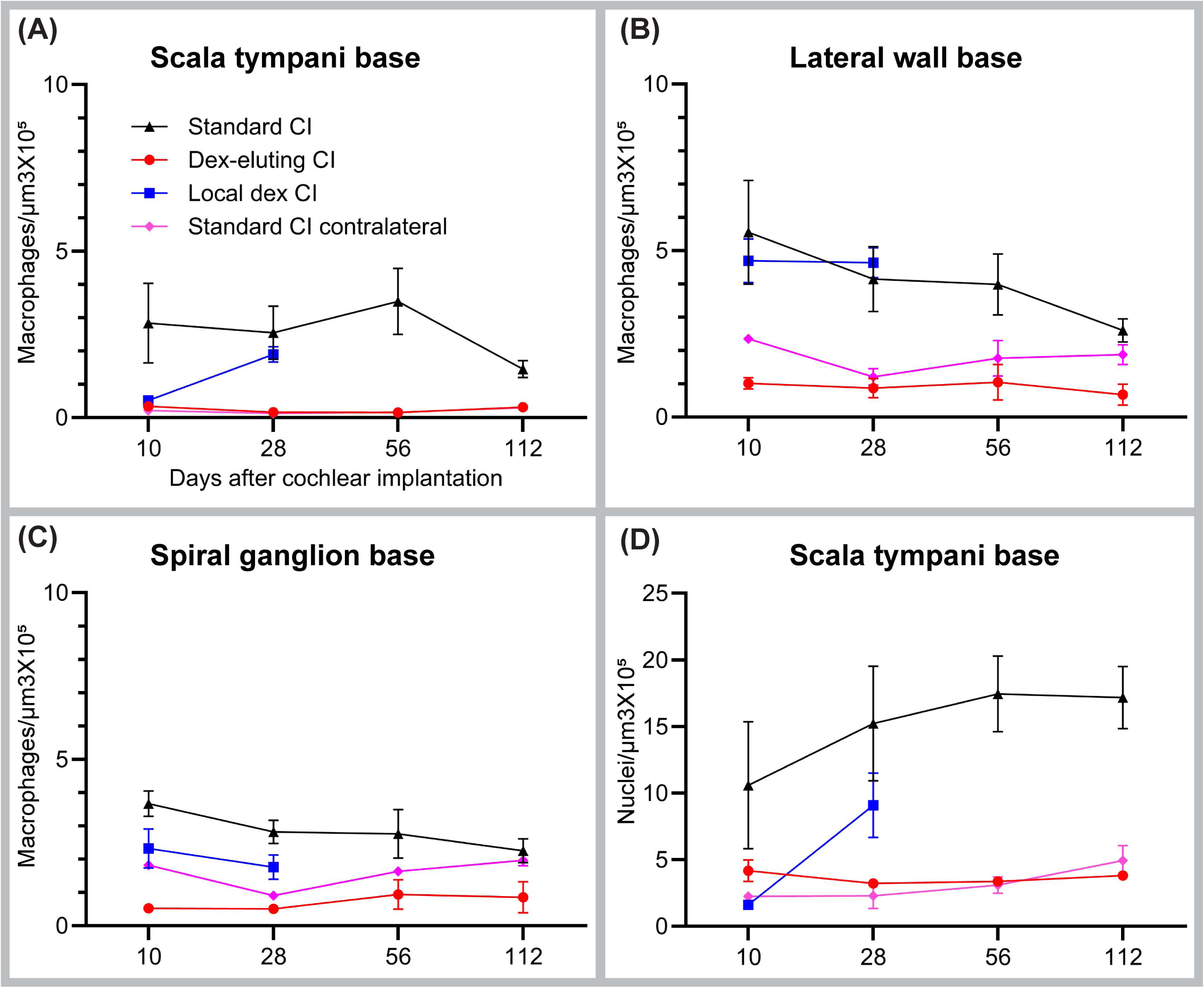
Quantification of macrophage and cellular density following cochlear implantation in basal cochlear turn Following tracing of the outline of scala tympani of the base of the cochlea, Rosenthal canal (RC), and lateral wall of the base of the cochlea, volumes were measured using IMARIS image analysis software. Using a supervised, automated counting system aided by a custom-made macro, counts of CX3CR1+ macrophages in each area were made and density was quantified. An average value from 3 sections from a cochlea was taken. Macrophage density in A. scala tympani B. Spiral ganglion and C. Lateral wall of the base of cochlea is shown. D. Nuclei labeled with Hoechst 3342 in 30-µm thick midmodiolar sections were quantified and density was calculated in traced area. Error bars indicate SEM. Statistical analysis was performed using two-way ANOVA with Tukey’s multiple comparisons. Cochlear implantation causes infiltration of CX3CR1+ macrophages in all three areas. Dexamethasone eluting implants reduce macrophage and cellular infiltration as long as 112 days post-CI. Local injection of dexamethasone reduces the CX3CR1+ macrophage and cellular infiltration at 10 days post-CI but not at 28 days post-CI (Multiple Mann-Whitney test)

In contrast to the dramatic effect of Dex-CI, Dex-local reduced macrophage density only in the scala tympani of the base of the cochlea (and not lateral wall or ganglion) at 10 days post-CI (p= 0.0181). This effect wore off 28 days post-CI (p=0.47) (multiple Mann-Whitney test).

### 3.4. Dexamethasone eluting cochlear implants reduces cellular density in scala tympani of the base of the cochlea for an extended period

An increase in the density of all nucleated cells (Hoechst+) was observed in the scala tympani at the base of the cochlea implanted with a standard CI, compared to the contralateral cochleae, as shown histologically in Figure 4 and quantified in Figure 5 (p=0.03, 0.0003, 0.0001 and 0.0001 at 10-, 28-, 56- and 112-days post-CI, respectively). Dexamethasone eluting cochlear implants significantly reduced the cell density in the scala tympani of the base of the cochlea throughout the experimental period. (p=0.0373, 0.0001, 0.0001, and 0.0001 at 10-, 28-, 56- and 112-days post-CI, respectively). (Figure 4 and Figure 5). In fact, despite the presence of the electrode array, Dex-CI nearly eliminated cellular infiltration into the basal turn of the scala tympani throughout the study period, mirroring the appearance of an unimplanted cochlea.

Dex-local reduced nucleus density only in the scala tympani of the base of the cochlea (and not lateral wall or ganglion) at 10 days post-CI (p= 0.0045); this effect wore off at 28 days post-CI (p=0.15) (Multiple Mann-Whitney test).

### 3.5 Dexamethasone eluting cochlear implants reduces α-SMA+ fibrotic response in cochlea

Following implantation of standard CI, within the scala tympani adjacent to the CI electrode array, α-SMA + fibrotic tissue growth was observed. (Fig 6 and Figure 7). Fibrosis was observed as early as 10 days post-CI and maintained throughout the period until 112-day post-CI (p=0.0001, 0.0001,0.0005 and 0.0007 at 10-, 28-, 56- and 112-days post-CI, respectively. As with cellular infiltration, Dex-CI dramatically reduced the α-SMA+ fibrotic tissue growth into the scala tympani of the base of the cochlea throughout the period we examined (p=0.0001, 0.0001, 0.0001 and 0.0021 for 10-, 28-, 56- and 112-days post-CI, respectively). (Figures 6 and 7). In contrast to Dex-CI, Dex-local did not reduce α-SMA+ fibrotic tissue growth into the scala tympani of the base of the cochlea (p=0.06) (Figures 6 and 7) (Multiple Mann-Whitney test).

**Figure 6:**
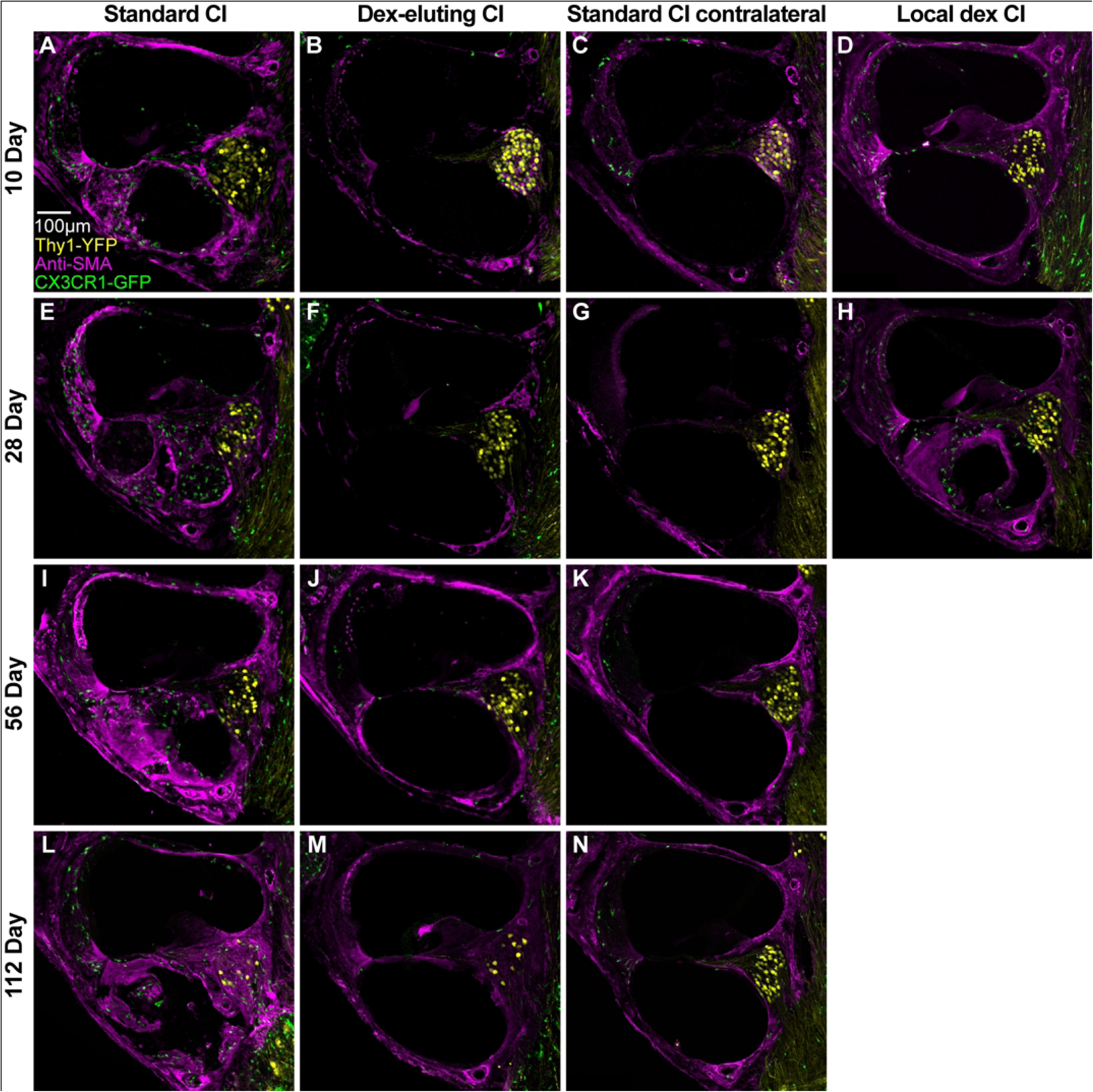
Dexamethasone eluting cochlear implants reduces α-SMA+ fibrotic tissue response following cochlear implantation. Following cochlear implantation and electrical stimulation, mice were euthanized at desired endpoints (10, 28, 56, or 112 days). Harvested cochleae were sectioned at 30-µm thickness, immunolabeled with anti-α-SMA antibody, and imaged with a confocal microscope. Scala tympani of the base of the cochlea was traced. The volumes of the scala tympani and the volume of the α-SMA+ fibrotic tissue were measured. Fibrosis was measured by dividing the α-SMA+ fibrotic tissue by the volume of the scala tympani and expressed in percentage. A-N. Representative images of midmodiolar sections labeled with anti-α-SMA antibody.

**Figure 7:**
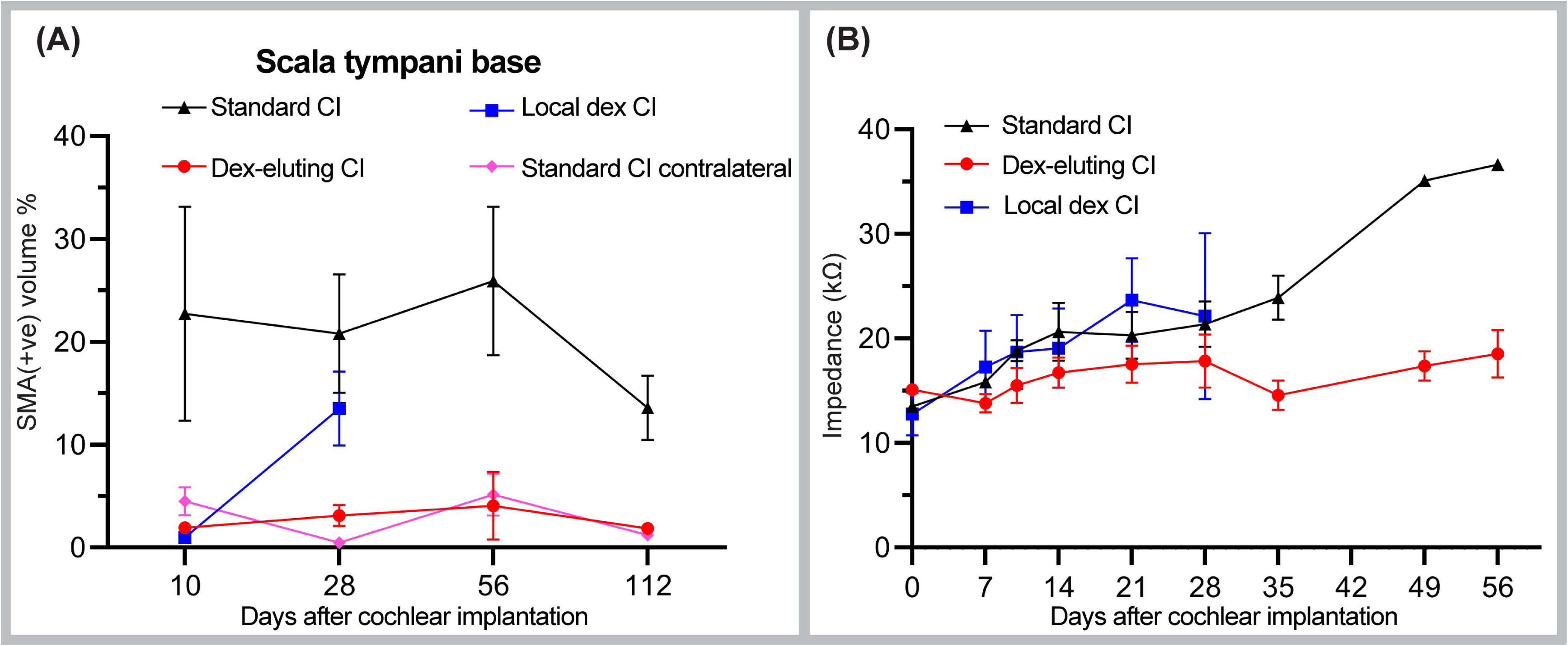
Quantification of α-SMA+ fibrotic tissue response and electrode impedance following cochlear implantation. A. Quantification of the α-SMA+ fibrotic tissue within scala tympani of the base of the cochlea. Error bars indicate SEM. Statistical analysis was performed using two-way ANOVA with Tukey’s multiple comparisons. Dexamethasone eluting cochlear implants reduce α-SMA+ fibrotic tissue response throughout the study period, i.e., 112 days post-CI. Local injection of dexamethasone reduces α-SMA+ fibrotic tissue response at 10 days post-CI but not at 28 days post-CI. B. Mean impedance values across functional electrodes at different time points are plotted. Error bars indicated SEM. Dexamethasone eluting implants reduce electrical impedance, Dex-local does not.

### 3.6 Dexamethasone Eluting cochlear implant reduces the electrical impedance of cochlear implants

Mean electrode impedance values for standard CI, Dex-CI, and Dex-local groups over time for active electrodes (electrodes with an open circuit denoting hardware failure were excluded) are shown in Figure 7. The trends in impedance growth over time for Standard, Dex-eluting, and Dex-local appear non-linear. Therefore, we used a linear mixed model using the square root of days, comparing the slope of the impedance over days between the standard CI, Dex-CI, and Dex-local. There is a significant interaction effect (p<.0001) which implies that the group effect changes over time.

Comparable baseline electrode impedance values were observed between the standard CI and Dex-CI (*p* =0.34,0) and standard-CI and Dex-local (0.99) groups at peri-operative baseline testing. Using contrasts (Kenward–Roger degrees of freedom and a Bonferroni alpha level correction) from the linear mixed model, we assessed at what point the three groups (Standard, Dex-CI, and Dex local) diverged in impedance growth over time. Dex-CI showed reduced electrode impedance compared to standard CI as early as 10 days post-CI (p=0.016) and continues to have lower impedance (p=0.0022, 0.0002, 0.0001, and 0.0001 at 14-, 21-, 28-, and 35-days post-CI). Dex-local implanted cochleae, on the other hand, had electrode impedances comparable to that of the standard CI (p=0.99 at 10, 14-, 21-, and 28-days post-CI). (Figure 7)

### 3.7 Dexamethasone eluting implants have a lower electrical impedance in human subjects

Figure 8 (A) summarizes the remote electrode impedance measurements (common ground mode). The means and standard deviations across all available remote checks for an individual are displayed. There is a clear separation across groups. Recipients of the standard 632 array tend to have higher electrode impedances. This tendency is more noticeable for basal and middle electrodes, compared to electrodes on the apical end of the array. The separation at the base partially reflects the frequent occurrence of non-current carrying electrodes. Clinical findings of non-auditory percepts or poor loudness growth are often addressed by deactivating the electrode in the patient’s MAP. Non-current-carrying electrodes tend to have higher electrical impedance values [85], which is more clearly observed in ears with standard 632 arrays but is not apparent in ears with 632D arrays. It is not clear why electrode impedances overlap across the two groups more at apical sites, especially given the apical location of the drug-eluting wells.

**Figure 8:**
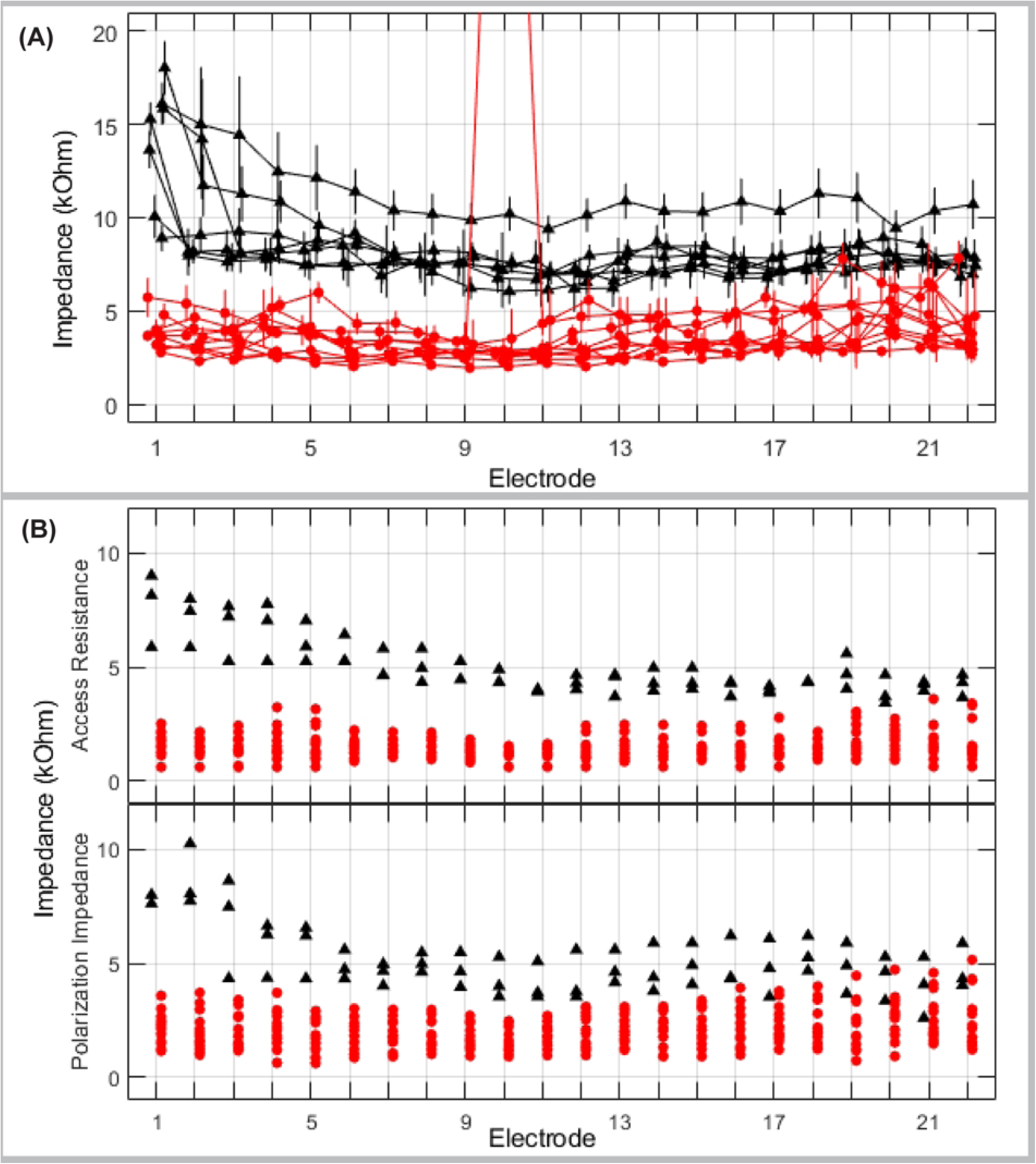
Impedance values from human subjects following cochlear implantation A. Summary of remotely measured impedance values from human subsects (1: most basal; 22: most apical electrode) for 17 participants. Black indicates a standard 632 array (N=7). Red indicates a 632D array (N=10). Triangles (632) or circles (632D) mark the mean across all available remote impedance measurements for a given participant; vertical bars extend +/- 1 standard deviation. Random jitter was added around the electrode number to help with visualization of individual data. B. Scatterplots for access resistance (top panel) and polarization impedance (bottom panel) at 3-months post activation to compare measurements in ears implanted with standard 632 arrays (black triangles; N=3) and ears with 632D arrays (red circles; N=15) for each electrode (1: most basal; 22: most apical).

The impedance data obtained at 3-months post activation are provided in Figure 8(B). Both access resistance and polarization impedance values tend to be higher for the recipients with standard 632 arrays; the separation between groups is largest at basal sites. Electrode deactivation is expected to impact the surface of the electrode, and thus would be reflected in the polarization component. The clear group polarization impedance separation for electrodes 1 and 2 suggests that dexamethasone keeps deposits from forming directly on the surface of the electrode, even when current is not being injected from that site. Access resistance is theoretically more sensitive to the conductivity of the tissue between the active and return path of current flow. Fibrotic tissue tends to increase over time in a base-to-apical fashion. [86] Although histological measures are not available for the human participants, the generally lower access resistance for the recipients of the 632D arrays, particularly at basal sites, is consistent with findings that dexamethasone reduces the formation of intracochlear fibrosis.

Despite the presumed release of a therapeutic dosage of dexamethasone ending around 1-month post-surgery, electrode impedance remains lower for the 632D recipients even at 3-months post activation. These results suggest a potentially long-lasting effect of dexamethasone beyond the presumed end of the release of significant levels of the drug.

## 4. Discussion

The data presented here demonstrate that diverse immune cells infiltrate cochlear tissues following implantation. Based on a reporter mouse model, macrophage infiltration continues for an extended period. This macrophage recruitment is associated with broad cellular infiltration and a fibrotic response and occurs universally in human and animal models of cochlear implantation. In our murine model, dexamethasone eluting cochlear implants reduced inflammatory and fibrotic response without affecting SGN survival. In human subjects, there was a reduction in electrical impedance.

Preclinical cochlear implant studies have predominantly used larger animal models (e.g. guinea pigs, sheep, gerbil, and cats. (Rahman, 2022 #1003) By contrast, mice have rarely been used, and even less so with active electrical stimulation, this is largely because of the technical difficulties of manufacturing and implanting functional electrode arrays and maintaining the electrical stimulation system in a small animal. Claussen et al. 2019 first described a mouse model of cochlear implantation with chronic electrical stimulation.[40] This was remarkable progress as various genetic tools (e.g. transgenic reporters, knockouts, RNA sequencing) are available for mouse models. Moreover, compared to any other animal models, the immune system of mice has been studied extensively. Leveraging the advantages of mouse models, Claussen et al. 2022 previously described that CX3CR1^+/GFP^ macrophage recruitment in implanted murine cochlea until 21 days post-implantation.[64] Subsequently, a study by Rahman et al 2023 revealed a sustained inflammatory and fibrotic response associated with rising electrode impedances until 56 days.

Rahman et al 2023 further observed that the depletion of macrophages did not reduce cellular infiltration or the extent of fibrotic response in the cochlea. Moreover, macrophage depletion increased electrode impedances and caused SGN degeneration. [65] <u>In</u> the current study leveraging the findings of Claussen et al. 2022 and Rahman et al. 2023, we extended our observations to 112-day post-CI. Moreover, we performed single-cell RNA sequencing to characterize the diversity of inflammatory cells and transcriptomic changes associated with cochlear implantation. Finally, we used a broad anti-inflammatory, immunosuppressive compound, dexamethasone (in two forms: a local injection that mirrors current clinical practice and dexamethasone eluting cochlear implant) to modulate the inflammatory FBR post-CI and its impact on electrical impedance and neural health.

A correlation between post-CI fibrous tissue growth and electrical impedance exists in animal models and dexamethasone eluting implants reduce both.[16]. Other studies have demonstrated the effectiveness of dexamethasone eluting implants in reducing fibrotic response. [87] Dexamethasone, a potent glucocorticoid, is known to suppress inflammation via various mechanisms.[88] In our study, dexamethasone eluting cochlear implants dramatically reduced macrophage density in implanted cochleae. However, macrophage depletion alone does not explain the reduction in overall cellular infiltration and fibrotic response caused by Dex-CIs since specific depletion of macrophages fails to mitigate scala tympani fibrosis following cochlear implantation, as previously observed.[65] Like the FBR elsewhere in the body[89], multiple immune cell types are associated with the FBR in an implanted cochlea: (T and B cells)[43], and eosinophils [90] have been reported in human cadaveric samples implanted with CI which is mirrored in our findings in implanted mouse cochleae. As in other parts of the body[89], cochlear implantation increases the expression of cytokines in implanted cochleae.[58] Dexamethasone inhibits T cell[91], B cell[92], eosinophils[93], and pro-inflammatory cytokines [94] while increasing expression of anti-inflammatory cytokines[95] providing a rational basis for its selection as the first drug to elute from electrode arrays. Nevertheless, as fibrosis can occur independently of inflammation[96, 97], the anti-fibrotic effect of Dex-CI could result from the modulation of pathways unrelated to inflammation.

Our current study presents some unique outcomes with important considerations. For the implant arrays used here, the dexamethasone is loaded into the Silicone carrier. Given the hydrophobic properties of Silicone, there are inherent limitations to the amount of drug that can be loaded into the Silicone. Further, elution from Silicone is predicted to occur relatively rapidly and there is limited inherent control to regulate the elution dynamics. [98] Nevertheless, our data support the long-term effectiveness of dexamethasone elution from Silicone of cochlear implant electrode arrays in suppressing tissue responses and elevation of electrode impedances. These findings might have different explanations: 1) An early burst release of dexamethasone suppressing the acute inflammatory response might be sufficient for long-term suppression of FBR. In this regard, it is relevant that the normal scala tympani is a fluid filled space and perhaps, if the acute inflammation is sufficiently suppressed, a long-term indwelling electrode array in the fluid filled scala does not generate a long-term FBR. 2) A very low rate of dexamethasone release from CI might be sufficient for the long-term suppression of FBR. 3) On the surface of biomaterials, adsorption of host proteins, complement activation, immune cell adhesion, and activation contribute to the FBR. [89] The localized presence of dexamethasone on the surface of the implant could potentially suppress FBR post-CI.

Local intratympanic administration of dexamethasone suspension is commonly used in clinical practice. However, the efficacy of locally administered dexamethasone has not been studied extensively. Some animal studies suggest that local dexamethasone reduces the inflammatory FBR following cochlear implantation. [99] while other animal studies suggest it is not effective. [100] Our study in the murine model is the first to show that local application of dexamethasone around the round window niche reduces the FBR initially; this effect appears to wear off after a few weeks. Our study raises the question of whether the current clinical practice, local dexamethasone, is effective in improving the long-term efficacy of cochlear implants. Further animal and human studies are required to answer this question. By contrast, the Dex-CIs used in this study dramatically reduce the FBR for an extended period.

Impedance data in our murine model and human subjects presented here also demonstrate that dexamethasone eluting implants reduce electrical impedance post-CI compared to standard CIs as seen in another recent study.[53] Total impedance is the composite of access resistance (determined by the resistance of the intracochlear environment) and polarization impedance (reflects capacitive resistive properties of the electrode-electrolyte interface)[19, 34, 101] Increased access resistance, observed in our human subjects implanted with standard CI, has been linked to FBR post-CI[17, 102] and loss of residual acoustic hearing[34]. In light of the findings in mice, reduction in access resistance in human subjects implanted with Dex-CI likely reflects reduced FBR; if so, Dex-CI also has the potential to prevent loss of residual acoustic hearing.

In this study, we have found that neither cochlear implantation nor dexamethasone eluting implants affect the SGN density post-CI. Previously, we have observed that in an implanted cochlea, depletion of macrophages with PLX5622 results in the degeneration of SGNs.[65] Like PLX5622, dexamethasone eluting implants also reduce macrophage density in the implanted cochlea. Why the dexamethasone eluting implant does not cause SGN degeneration despite reducing macrophage density in the cochlea is a question to be answered. One possible explanation is that macrophages play a protective role for SGNs in cochlea while other inflammatory cells and cytokines have a neurotoxic role. Some studies have shown a neuroprotective role of cochlear macrophages. [103–105] On the other hand, non-specific immunosuppressive drugs like dexamethasone and ibuprofen have been shown to protect SGN survival. [106] As a broad anti-inflammatory and immunosuppressive drug, dexamethasone can potentially suppress the neurotoxic inflammatory cells and cytokines while PLX5622 only suppresses the neuroprotective macrophages. While, as shown here and elsewhere, a diversity of immune and inflammatory cells infiltrate the cochlea after implantation, understanding the impact of these non-macrophage immune cells on CI outcomes requires further investigation.

In summary, the data presented here demonstrate the remarkable effectiveness of dexamethasone eluting electrode arrays to suppress the universal inflammatory FBR post-CI. Multiple clinical trials (NCT06142682, NCT04750642, NCT06424262) are ongoing on dexamethasone eluting cochlear implants focused on speech intelligibility, hearing preservation, and electrode impedances. The remarkable effectiveness of dexamethasone eluting cochlear implants in murine models as well as human subjects to suppress the long-term intracochlear tissue responses to implanted electrode arrays can guide future clinical trials and translational research that stand to improve the functional outcomes of cochlear implantation.

## Supporting information

Supplementary Figure S1: Quality Control (QC) metrics for single cell RNA-Seq of CD11b+ immune cells.

Supplementary Figure S2: Quantification of macrophage infiltration following cochlear implantation in the middle, apical cochlear turns

Supplementary Figure S3: Quantification of cellular density in the spiral ganglia, lateral walls of Cochlea following implantation

Supplementary Figure S4: CX3CR1+ MHCII+ macrophages in implanted cochlea

Supplementary Figure S5: Quantification of CX3CR1+ MHCII+ macrophages in implanted cochlea

Supplementary Figure S6: Spiral ganglion neurons (SGN) density following cochlear implantation

## Data Availability

All data produced in the present study are available upon reasonable request to the authors.

## Acknowledgements

The authors thank Kapila de Silva (Cochlear Limited) for his technical support for UPLC experiments for dexamethasone eluting implants from the murine model. The authors are grateful to Amy Bussa (Department of Otolaryngology-Head and Neck Surgery, The University of Iowa, Iowa City, IA, United States) for her dependability in sending notifications and email reminders to subjects in regard to the daily remote impedance measurements. Authors would also like to acknowledge Madeline Pyle (Auditory Development and Restoration Program, Neurotology Branch, *National Institute on Deafness and Other Communication Disorders, National Institutes of Health, Bethesda, MD, United States*) for her work in streamlining our immune cell scRNA-Seq protocol.

## References

1. Olusanya, B.O., A.C. Davis, and H.J. Hoffman, Hearing loss: rising prevalence and impact. Bull World Health Organ, 2019. 97(10): p. 646–646A.

2. Ching, T.Y., et al., Language development and everyday functioning of children with hearing loss assessed at 3 years of age. Int J Speech Lang Pathol, 2010. 12(2): p. 124–31.

3. Ciorba, A., et al., The impact of hearing loss on the quality of life of elderly adults. Clin Interv Aging, 2012. 7: p. 159–63.

4. Elbeltagy, R., Prevalence of Mild Hearing Loss in Schoolchildren and its Association with their School Performance. Int Arch Otorhinolaryngol, 2020. 24(1): p. e93–e98.

5. Livingston, G., et al., Dementia prevention, intervention, and care: 2020 report of the Lancet Commission. Lancet, 2020. 396(10248): p. 413–446.

6. Livingston, G., et al., Dementia prevention, intervention, and care. Lancet, 2017. 390(10113): p. 2673–2734.

7. Powell, D.S., et al., Hearing Loss and Cognition: What We Know and Where We Need to Go. Front Aging Neurosci, 2021. 13: p. 769405.

8. Cheng, A.G., L.L. Cunningham, and E.W. Rubel, Mechanisms of hair cell death and protection. Curr Opin Otolaryngol Head Neck Surg, 2005. 13(6): p. 343–8.

9. Mudry, A. and M. Mills, The early history of the cochlear implant: a retrospective. JAMA Otolaryngol Head Neck Surg, 2013. 139(5): p. 446–53.

10. Roche, J.P. and M.R. Hansen, On the Horizon: Cochlear Implant Technology. Otolaryngol Clin North Am, 2015. 48(6): p. 1097–116.

11. Jensen, M.J., et al., Cochlear implant material effects on inflammatory cell function and foreign body response. Hear Res, 2022. 426: p. 108597.

12. Buchman, C.A., et al., Assessment of Speech Understanding After Cochlear Implantation in Adult Hearing Aid Users: A Nonrandomized Controlled Trial. JAMA Otolaryngol Head Neck Surg, 2020. 146(10): p. 916–924.

13. Clark, G.M., et al., Cochlear implantation: osteoneogenesis, electrode-tissue impedance, and residual hearing. Ann Otol Rhinol Laryngol Suppl, 1995. 166: p. 40–2.

14. Ni, D., et al., Cochlear pathology following chronic electrical stimulation of the auditory nerve. I: Normal hearing kittens. Hear Res, 1992. 62(1): p. 63–81.

15. Shepherd, R.K., et al., Cochlear pathology following reimplantation of a multichannel scala tympani electrode array in the macaque. Am J Otol, 1995. 16(2): p. 186–99.

16. Wilk, M., et al., Impedance Changes and Fibrous Tissue Growth after Cochlear Implantation Are Correlated and Can Be Reduced Using a Dexamethasone Eluting Electrode. PLoS One, 2016. 11(2): p. e0147552.

17. Xu, J., et al., Chronic electrical stimulation of the auditory nerve at high stimulus rates: a physiological and histopathological study. Hear Res, 1997. 105(1-2): p. 1–29.

18. Shaul, C., et al., Electrical Impedance as a Biomarker for Inner Ear Pathology Following Lateral Wall and Peri-modiolar Cochlear Implantation. Otol Neurotol, 2019. 40(5): p. e518–e526.

19. Tykocinski, M., L.T. Cohen, and R.S. Cowan, Measurement and analysis of access resistance and polarization impedance in cochlear implant recipients. Otol Neurotol, 2005. 26(5): p. 948–56.

20. Kirk, J.R., D. Smyth, and W.F. Dueck, A new paradigm of hearing loss and preservation with cochlear implants: Learnings from fundamental studies and clinical research. Hear Res, 2023. 433: p. 108769.

21. Shim, H., et al., Differences in neural encoding of speech in noise between cochlear implant users with and without preserved acoustic hearing. Hear Res, 2023. 427: p. 108649.

22. Tarabichi, O., M. Jensen, and M.R. Hansen, Advances in hearing preservation in cochlear implant surgery. Curr Opin Otolaryngol Head Neck Surg, 2021. 29(5): p. 385–390.

23. Gantz, B.J., M. Hansen, and C.C. Dunn, Clinical perspective on hearing preservation in cochlear implantation, the University of Iowa experience. Hear Res, 2022. 426: p. 108487.

24. Kelsall, D.C., R.J.G. Arnold, and L. Lionnet, Patient-Reported Outcomes From the United States Clinical Trial for a Hybrid Cochlear Implant. Otol Neurotol, 2017. 38(9): p. 1251–1261.

25. Li, C., M. Kuhlmey, and A.H. Kim, Electroacoustic Stimulation. Otolaryngol Clin North Am, 2019. 52(2): p. 311–322.

26. Wolfe, J., et al., Potential Benefits of an Integrated Electric-Acoustic Sound Processor with Children: A Preliminary Report. J Am Acad Audiol, 2017. 28(2): p. 127–140.

27. Woodson, E.A., et al., The Hybrid cochlear implant: a review. Adv Otorhinolaryngol, 2010. 67: p. 125–134.

28. O’Leary, S.J., et al., Relations between cochlear histopathology and hearing loss in experimental cochlear implantation. Hear Res, 2013. 298: p. 27–35.

29. O’Malley, J.T., et al., Delayed hearing loss after cochlear implantation: Re-evaluating the role of hair cell degeneration. Hear Res, 2024. 447: p. 109024.

30. Quesnel, A.M., et al., Delayed loss of hearing after hearing preservation cochlear implantation: Human temporal bone pathology and implications for etiology. Hear Res, 2016. 333: p. 225–234.

31. Scheperle, R.A., et al., Delayed changes in auditory status in cochlear implant users with preserved acoustic hearing. Hear Res, 2017. 350: p. 45–57.

32. Schraivogel, S., et al., Cochlear implant electrode impedance subcomponents as biomarker for residual hearing. Front Neurol, 2023. 14: p. 1183116.

33. Shepherd, R.K., et al., Cochlear pathology following chronic electrical stimulation using non charge balanced stimuli. Acta Otolaryngol, 1991. 111(5): p. 848–60.

34. Tejani, V.D., et al., Access and Polarization Electrode Impedance Changes in Electric-Acoustic Stimulation Cochlear Implant Users with Delayed Loss of Acoustic Hearing. J Assoc Res Otolaryngol, 2022. 23(1): p. 95–118.

35. Wimmer, W., et al., Cochlear Implant Electrode Impedance as Potential Biomarker for Residual Hearing. Front Neurol, 2022. 13: p. 886171.

36. Zhang, H., G. Stark, and L. Reiss, Changes in Gene Expression and Hearing Thresholds After Cochlear Implantation. Otol Neurotol, 2015. 36(7): p. 1157–65.

37. Noonan, K.Y., et al., Immune Response of Macrophage Population to Cochlear Implantation: Cochlea Immune Cells. Otol Neurotol, 2020. 41(9): p. 1288–1295.

38. Okayasu, T., et al., The Distribution and Prevalence of Macrophages in the Cochlea Following Cochlear Implantation in the Human: An Immunohistochemical Study Using Anti-Iba1 Antibody. Otol Neurotol, 2020. 41(3): p. e304–e316.

39. O’Malley, J.T., J.B. Nadol, Jr., and M.J. McKenna, Anti CD163+, Iba1+, and CD68+ Cells in the Adult Human Inner Ear: Normal Distribution of an Unappreciated Class of Macrophages/Microglia and Implications for Inflammatory Otopathology in Humans. Otol Neurotol, 2016. 37(1): p. 99–108.

40. Claussen, A.D., et al., A mouse model of cochlear implantation with chronic electric stimulation. PLoS One, 2019. 14(4): p. e0215407.

41. Irving, S., et al., Cochlear implantation for chronic electrical stimulation in the mouse. Hear Res, 2013. 306: p. 37–45.

42. Mistry, N., et al., Cochlear implantation in the mouse via the round window: effects of array insertion. Hear Res, 2014. 312: p. 81–90.

43. Nadol, J.B., Jr., et al., Cellular immunologic responses to cochlear implantation in the human. Hear Res, 2014. 318: p. 11–7.

44. Bas, E., et al., Spiral ganglion cells and macrophages initiate neuro-inflammation and scarring following cochlear implantation. Front Cell Neurosci, 2015. 9: p. 303.

45. Hinz, B., et al., Alpha-smooth muscle actin expression upregulates fibroblast contractile activity. Mol Biol Cell, 2001. 12(9): p. 2730–41.

46. Trojanowska, M., et al., Pathogenesis of fibrosis: type 1 collagen and the skin. J Mol Med (Berl), 1998. 76(3-4): p. 266–74.

47. Fleet, A., et al., Outcomes following cochlear implantation with eluting electrodes: A systematic review. Laryngoscope Investig Otolaryngol, 2024. 9(3): p. e1263.

48. Rahman, M.T., et al., Cochlear implants: Causes, effects and mitigation strategies for the foreign body response and inflammation. Hear Res, 2022. 422: p. 108536.

49. Ardic, F.N., et al., The Effect of Intracochlear and Intratympanic Dexamethasone on Cochlear Implant Impedance. Turk Arch Otorhinolaryngol, 2023. 61(3): p. 103–108.

50. Lo, J., et al., The Role of Preoperative Steroids in Atraumatic Cochlear Implantation Surgery. Otol Neurotol, 2017. 38(8): p. 1118–1124.

51. James, D.P., et al., Effects of round window dexamethasone on residual hearing in a Guinea pig model of cochlear implantation. Audiol Neurootol, 2008. 13(2): p. 86–96.

52. Manrique-Huarte, R., et al., Cochlear Implantation With a Dexamethasone Eluting Electrode Array: Functional and Anatomical Changes in Non-Human Primates. Otol Neurotol, 2020. 41(7): p. e812–e822.

53. Briggs, R., et al., Comparison of electrode impedance measures between a dexamethasone-eluting and standard Cochlear Contour Advance(R) electrode in adult cochlear implant recipients. Hear Res, 2020. 390: p. 107924.

54. Ahmadi, N., et al., Long-term effects and potential limits of intratympanic dexamethasone-loaded hydrogels combined with dexamethasone-eluting cochlear electrodes in a low-insertion trauma Guinea pig model. Hear Res, 2019. 384: p. 107825.

55. Bas, E., et al., Laminin-coated electrodes improve cochlear implant function and post-insertion neuronal survival. Neuroscience, 2019. 410: p. 97–107.

56. Bas, E., et al., Electrode array-eluted dexamethasone protects against electrode insertion trauma induced hearing and hair cell losses, damage to neural elements, increases in impedance and fibrosis: A dose response study. Hear Res, 2016. 337: p. 12–24.

57. Liu, Y., et al., Effects of a dexamethasone-releasing implant on cochleae: A functional, morphological and pharmacokinetic study. Hear Res, 2015. 327: p. 89–101.

58. Simoni, E., et al., Immune Response After Cochlear Implantation. Front Neurol, 2020. 11: p. 341.

59. Van De Water, T.R., et al., Conservation of hearing and protection of auditory hair cells against trauma-induced losses by local dexamethasone therapy: molecular and genetic mechanisms. Cochlear Implants Int, 2010. 11 **Suppl 1**: p. 42–55.

60. Feng, G., et al., Imaging neuronal subsets in transgenic mice expressing multiple spectral variants of GFP. Neuron, 2000. 28(1): p. 41–51.

61. Jung, S., et al., Analysis of fractalkine receptor CX(3)CR1 function by targeted deletion and green fluorescent protein reporter gene insertion. Mol Cell Biol, 2000. 20(11): p. 4106–14.

62. Reiss, L.A.J., et al., Animal Models of Hearing Loss after Cochlear Implantation and Electrical Stimulation. Hear Res, 2022. 426: p. 108624.

63. Scheper, V., et al., Randomized placebo-controlled clinical trial investigating the effect of antioxidants and a vasodilator on overall safety and residual hearing preservation in cochlear implant patients. Trials, 2020. 21(1): p. 643.

64. Claussen, A.D., et al., Chronic cochlear implantation with and without electric stimulation in a mouse model induces robust cochlear influx of CX3CR1(+/GFP) macrophages. Hear Res, 2022. 426: p. 108510.

65. Rahman, M.T., et al., Contribution of macrophages to neural survival and intracochlear tissue remodeling responses following cochlear implantation. J Neuroinflammation, 2023. 20(1): p. 266.

66. Yamahara, K., et al., Hearing preservation at low frequencies by insulin-like growth factor 1 in a guinea pig model of cochlear implantation. Hear Res, 2018. 368: p. 92–108.

67. Korrapati, S., et al., Single Cell and Single Nucleus RNA-Seq Reveal Cellular Heterogeneity and Homeostatic Regulatory Networks in Adult Mouse Stria Vascularis. Front Mol Neurosci, 2019. 12: p. 316.

68. Wolock, S.L., R. Lopez, and A.M. Klein, Scrublet: Computational Identification of Cell Doublets in Single-Cell Transcriptomic Data. Cell Syst, 2019. 8(4): p. 281–291 e9.

69. Kopelovich, J.C., et al., Intracochlear electrical stimulation suppresses apoptotic signaling in rat spiral ganglion neurons after deafening in vivo. Otolaryngol Head Neck Surg, 2013. 149(5): p. 745–52.

70. Alexander D. Claussen, R.V.Q., Timon Higgins, Brian Mostaert, Muhammad Taifur Rahman, Jonathon Kirk, Keiko Hirose, Marlan R. Hansen, Chronic Cochlear Implantation with and without Electric Stimulation in a Mouse Model Induces Robust Cochlear Influx of CX3CR1+/GFP Macrophages, T.U.o. Iowa, Editor. 2021, Biorxiv.

71. Ali, S., et al., Functional genetic variation in NFKBIA and susceptibility to childhood asthma, bronchiolitis, and bronchopulmonary dysplasia. J Immunol, 2013. 190(8): p. 3949–58.

72. Cartwright, T., N.D. Perkins, and L.W. C, NFKB1: a suppressor of inflammation, ageing and cancer. FEBS J, 2016. 283(10): p. 1812–22.

73. Nurmi, K., et al., Truncating NFKB1 variants cause combined NLRP3 inflammasome activation and type I interferon signaling and predispose to necrotizing fasciitis. Cell Rep Med, 2024. 5(4): p. 101503.

74. Zhang, G.L., et al., Association of the NFKBIA gene polymorphisms with susceptibility to autoimmune and inflammatory diseases: a meta-analysis. Inflamm Res, 2011. 60(1): p. 11–8.

75. Aouadi, M., et al., Orally delivered siRNA targeting macrophage Map4k4 suppresses systemic inflammation. Nature, 2009. 458(7242): p. 1180–4.

76. Chuang, H.C., X. Wang, and T.H. Tan, MAP4K Family Kinases in Immunity and Inflammation. Adv Immunol, 2016. 129: p. 277–314.

77. Meng, B., et al., Myeloid-derived growth factor inhibits inflammation and alleviates endothelial injury and atherosclerosis in mice. Sci Adv, 2021. 7(21).

78. Virbasius, J.V. and M.P. Czech, Map4k4 Signaling Nodes in Metabolic and Cardiovascular Diseases. Trends Endocrinol Metab, 2016. 27(7): p. 484–492.

79. Choi, S., et al., Proton pump inhibitors exert anti-allergic effects by reducing TCTP secretion. PLoS One, 2009. 4(6): p. e5732.

80. Kim, Y.H., J.R. Lee, and M.J. Hahn, Regulation of inflammatory gene expression in macrophages by epithelial-stromal interaction 1 (Epsti1). Biochem Biophys Res Commun, 2018. 496(2): p. 778–783.

81. Zhang, J., et al., S100A4 promotes colon inflammation and colitis-associated colon tumorigenesis. Oncoimmunology, 2018. 7(8): p. e1461301.

82. Miyake, K., et al., Single-cell transcriptomics identifies the differentiation trajectory from inflammatory monocytes to pro-resolving macrophages in a mouse skin allergy model. Nat Commun, 2024. 15(1): p. 1666.

83. Waddell, L.A., et al., ADGRE1 (EMR1, F4/80) Is a Rapidly-Evolving Gene Expressed in Mammalian Monocyte-Macrophages. Front Immunol, 2018. 9: p. 2246.

84. Lu, Y. and Z. Jia, Inflammation-Related Gene Signature for Predicting the Prognosis of Head and Neck Squamous Cell Carcinoma. Int J Gen Med, 2022. 15: p. 4793–4805.

85. Dorman, M.F., et al., The coding of vowel identity by patients who use the Ineraid cochlear implant. J Acoust Soc Am, 1992. 92(6): p. 3428–31.

86. Nadol, J.B., Jr., et al., Histopathology of cochlear implants in humans. Ann Otol Rhinol Laryngol, 2001. 110(9): p. 883–91.

87. Toulemonde, P., et al., Evaluation of the Efficacy of Dexamethasone-Eluting Electrode Array on the Post-Implant Cochlear Fibrotic Reaction by Three-Dimensional Immunofluorescence Analysis in Mongolian Gerbil Cochlea. J Clin Med, 2021. 10(15).

88. Barnes, P.J., Anti-inflammatory actions of glucocorticoids: molecular mechanisms. Clin Sci (Lond), 1998. 94(6): p. 557–72.

89. Anderson, J.M., A. Rodriguez, and D.T. Chang, Foreign body reaction to biomaterials. Semin Immunol, 2008. 20(2): p. 86–100.

90. Seyyedi, M. and J.B. Nadol, Jr., Intracochlear inflammatory response to cochlear implant electrodes in humans. Otol Neurotol, 2014. 35(9): p. 1545–51.

91. Giles, A.J., et al., Dexamethasone-induced immunosuppression: mechanisms and implications for immunotherapy. J Immunother Cancer, 2018. 6(1): p. 51.

92. Andreau, K., et al., Induction of apoptosis by dexamethasone in the B cell lineage. Immunopharmacology, 1998. 40(1): p. 67–76.

93. Schleimer, R.P. and B.S. Bochner, The effects of glucocorticoids on human eosinophils. J Allergy Clin Immunol, 1994. 94(6 Pt 2): p. 1202–13.

94. Nehme, A. and J. Edelman, Dexamethasone inhibits high glucose-, TNF-alpha-, and IL-1beta-induced secretion of inflammatory and angiogenic mediators from retinal microvascular pericytes. Invest Ophthalmol Vis Sci, 2008. 49(5): p. 2030–8.

95. Agarwal, S.K. and G.D. Marshall, Jr., Dexamethasone promotes type 2 cytokine production primarily through inhibition of type 1 cytokines. J Interferon Cytokine Res, 2001. 21(3): p. 147–55.

96. Mack, M., Inflammation and fibrosis. Matrix Biol, 2018. 68-69: p. 106-121.

97. Wynn, T.A., Cellular and molecular mechanisms of fibrosis. J Pathol, 2008. 214(2): p. 199–210.

98. Cauldbeck, H., et al., Controlling drug release from non-aqueous environments: Moderating delivery from ocular silicone oil drug reservoirs to combat proliferative vitreoretinopathy. J Control Release, 2016. 244(Pt A): p. 41–51.

99. Lyu, A.R., et al., Effects of dexamethasone on intracochlear inflammation and residual hearing after cochleostomy: A comparison of administration routes. PLoS One, 2018. 13(3): p. e0195230.

100. Lee, J., et al., Effect of both local and systemically administered dexamethasone on long-term hearing and tissue response in a Guinea pig model of cochlear implantation. Audiol Neurootol, 2013. 18(6): p. 392–405.

101. Dymond, A.M., Characteristics of the metal-tissue interface of stimulation electrodes. IEEE Trans Biomed Eng, 1976. 23(4): p. 274–80.

102. Tykocinski, M., et al., Chronic electrical stimulation of the auditory nerve using high surface area (HiQ) platinum electrodes. Hear Res, 2001. 159(1-2): p. 53–68.

103. Kaur, T., et al., Fractalkine Signaling Regulates Macrophage Recruitment into the Cochlea and Promotes the Survival of Spiral Ganglion Neurons after Selective Hair Cell Lesion. J Neurosci, 2015. 35(45): p. 15050–61.

104. Manickam, V., et al., Macrophages Promote Repair of Inner Hair Cell Ribbon Synapses following Noise-Induced Cochlear Synaptopathy. J Neurosci, 2023. 43(12): p. 2075–2089.

105. Yagihashi, A., T. Sekiya, and S. Suzuki, Macrophage colony stimulating factor (M-CSF) protects spiral ganglion neurons following auditory nerve injury: morphological and functional evidence. Exp Neurol, 2005. 192(1): p. 167–77.

106. Rahman, M.T., et al., Anti-inflammatory Therapy Protects Spiral Ganglion Neurons After Aminoglycoside Antibiotic-Induced Hair Cell Loss. Neurotherapeutics, 2023. 20(2): p. 578–601.

107. Hirayama, D., T. Iida, and H. Nakase, The Phagocytic Function of Macrophage-Enforcing Innate Immunity and Tissue Homeostasis. Int J Mol Sci, 2017. 19(1).

108. Roche, P.A. and K. Furuta, The ins and outs of MHC class II-mediated antigen processing and presentation. Nat Rev Immunol, 2015. 15(4): p. 203–16.

