## Supplementary data for "Cochlear implants with dexamethasone-eluting electrode arrays reduce foreign body response in a murine model of cochlear implantation and human subjects"

**Supplementary Information:**

**Macrophage density in middle and apical turns:**

In the middle cochlear turn, a higher density of CX3CR1+macrophages in the spiral ganglia was observed in the standard CI group at 56 days post-CI (p=0.026) and returned to a state comparable to the contralateral unimplanted cochlea by 112 days post-CI(p=0.98). Dex-CI reduces the CX3CR1+macrophages in the spiral ganglia of the middle turn throughout the time examined (p=0.0001, 0.009, 0.0025, 0.005 at 10-, 28-, 56-, and 112-days post-CI).

In the lateral wall of the middle cochlear turn, a higher density of CX3CR1+ macrophages is observed in the standard CI group at 28 days (p= 0.029) and 56 days (p=0.014) post-CI. By 112 days post-CI, CX3CR1+macrophage density is comparable to the contralateral cochlea (p=0.93). Dex-CI reduced the CX3CR1+macrophage density until 56 days post-CI (p=0.0001, 0.0116, 0.001 at 10-, 28-, and 56-days post-CI).

In the apical cochlear turn, a higher density of CX3CR1+ macrophages is observed in the standard CI group at 10 days post-CI (p=0.0013) Dex-CI keeps the macrophage density lower than the standard CI group until 56 days post-CI (p=0.0013, 0.069, 0.008 at 10-, 28-, and 56-days post-CI). In the lateral of the apical turn, no change in macrophage density was observed after standard CI implantation at any time point examined. A reduction in macrophage density was evident at 10-day post-CI (p=0.0001) Statistical analysis was performed using two-way ANOVA with Tukey's multiple comparisons. (Figure 4 and Supplementary Figure S2)

There was no significant effect of Dex-local on the macrophage density in the spiral ganglia and lateral wall of middle and apical turns of the cochlea (p>0.05) (Multiple Mann-Whitney test).

**Nucleus density in spiral ganglia, and lateral walls:**

Standard CI does not cause cellular infiltration in areas other than ST of the base of cochlea ($p>0.05$ for all locations) Dex-CI (two-way ANOVA with Tukey's multiple comparisons) or Dex-local (Multiple Mann-Whitney test) do not affect the cellular density in other areas of the cochlea ($p>0.05$) (Figure 4 and Supplementary Figure S3)

**Dexamethasone eluting implants suppress MHCII expression on CX3CR1+ macrophage in the cochlea following cochlear implantation in murine model:**

Macrophages have a critical role in innate immunity and tissue homeostasis. [107] Additionally, macrophages are known to express antigen-presenting molecule, MHCII. MHCII-mediated antigen presentation to CD4+T cells is a key mechanism for adaptive immune response. [108] In this study, we examined the effects of cochlear implantation on MHCII expression in CX3CR1-positive macrophages.

In the scala tympani of the base of the cochlea, compared to the contralateral unimplanted cochlea, a delayed (112-days post-CI) increase in CX3CR1+MHCII+ macrophage density was noticed ($p=0.0022$) while there was no recruitment of CX3CR1+MHCII+ macrophage at 10 days post-CI ($p=0.052$).

Dexamethasone eluting cochlear implants reduces the density of CX3CR1+MHCII+ macrophages throughout the cochlea for an extended (112 days) period ($p<0.05$ for scala tympani, spiral ganglia, and lateral walls of the base of the cochlea for 10, 28, 56 and 112 days). Dex-Local does not affect CX3CR1+MHCII+ macrophage density ($p>0.05$ for scala tympani, spiral ganglia, and lateral walls of the base of the cochlea at 10- and 28-days post-CI) (Supplementary Figure S4 and S5). Statistical analysis performed with Multiple Mann-Whitney test.

**Dexamethasone eluting implants do not affect SGN density following implantation:**

Supplementary Figure S7 shows the representative images of Thy1$^{YFP}$+ SGNs from implanted and contralateral cochlea from 'Standard CI' and implanted cochlea from 'Dex-CI' group and the statistical analyses of the data. We observed SGN degeneration in middle cochlear turn (p=0.048) and a trend for the base of the cochlea (p=0.12) compared to respective contralateral, unimplanted groups at 112 days post-CI. Dexamethasone eluting implants do not affect the SGN density (p=0.99, 0.99, and 0.99, for basal, middle, and apical turns, respectively. Statistical analyses with two-way ANOVA with Tukey multiple comparisons.)

**Supplementary Figures**

**Supplementary Figure S1:** Quality Control (QC) metrics for single cell RNA-Seq of CD11b+ immune cells. A, Box-and-whisker plots demonstrates distribution of genes as determined by counts in CD11b+-immune cells isolated from control (Ctrl) and implanted cochlea. Bottom of box is the lower quartile, followed by median, followed by the top of the box, representing the upper quartile. The box represents the interquartile range (IQR). Top and bottom whiskers represent the upper and lower extreme, respectively.  B, Similarly, box-and-whisker plots demonstrates total count distribution between Ctrl and implanted cochlea. C, Box-and-whisker plots demonstrate mitochondrial percentage distribution in CD11b+-immune cells isolated from Ctrl and implanted cochlea. Mitochondrial percentage was less than 5% in both samples. D, Finally, starting cell counts and ending cell counts after filtering steps are shown in the bar graph with the last red bar indicative of the final count number after application of Dedoublet.

**Supplementary Figure S2: Quantification of macrophage infiltration following cochlear implantation in the middle, apical cochlear turns**

Maximum intensity z-projections of 3D confocal image stacks of 30-µm thick midmodiolar sections were used to count CX3CR1+ macrophage cells using IMARIS image analysis software. Following tracing of the outline of the Rosenthal canal (RC) and lateral wall middle and apical turn of the cochlea, volumes were measured. Using a supervised, automated counting system aided by a custom-made macro, counts of CX3CR1+ macrophages were done. density was quantified in each area measured. An average value from 3 sections from a cochlea was taken. Macrophage density in A. Spiral ganglion of the middle turn, B. Lateral wall of the middle turn C. B. Spiral ganglion of apex, and D. Lateral wall of the apical turn of the cochlea is shown. Error bars indicate SEM. Dexamethasone eluting implants, reduce macrophage infiltration. Statistical analysis was performed using two-way ANOVA with Tukey's multiple comparisons and the Multiple Mann-Whitney test.

**Supplementary Figure S3: Quantification of cellular density in the spiral ganglia, lateral walls of Cochlea following implantation**

Following cochlear implantation and electrical stimulation, mice were euthanized at desired time points (10, 28, 56, or 112 days) Nuclei labeled with Hoechst 3342 in 30-μm thick midmodiolar sections were quantified and density calculated in traced area. Error bars indicate SEM. Statistical analysis was performed using two-way ANOVA with Tukey's multiple comparisons and the Multiple Mann-Whitney test.

**Supplementary Figure S4: CX3CR1+ MHCII+ macrophages in implanted cochlea**

30-μm thick midmodiolar sections were labeled with anti-MHCII antibody and imaged. Maximum intensity z-projections of 3D confocal image stacks were used to count CX3CR1+MHCII+ macrophage cells. Fluorescent microscopic images of representative mid-modiolar sections of the basal turn of the cochlea are shown.

**Supplementary Figure S5: Quantification of CX3CR1+ MHCII+ macrophages in implanted cochlea**

30-μm thick midmodiolar sections labeled with MHCII antibody were analyzed using IMARIS image analysis software. Scala tympani of the base and Rosenthal canal (RC) and lateral wall of the base, middle and apical turn of the cochlea were traced and volumes were measured. Using a supervised, automated counting system aided by a custom-made macro, counts of

CX3CR1+MHCII+ macrophages were made. In every area traced, CX3CR1+MHCII+ macrophage density was quantified. An average value from 3 sections from a cochlea was taken. Error bars indicate SEM. Statistical analyses were done using the Multiple Mann-Whitney test.

**Supplementary Figure S6: Spiral ganglion neurons (SGN) density following cochlear implantation.**

Following cochlear implantation and electrical stimulation, cochleae were harvested at desired endpoints (10, 28, 56, or 112 days). 30-μm thick midmodiolar sections from the implanted $CX3CR1^{+/GFP}$ $Thy1^{+/YFP}$ were imaged. Representative images from the spiral ganglion of the base of the cochlea from A. Standard CI B. Dex-CI and C. Contralateral to standard CI. Rosenthal canals were manually traced, and Thy1+ SGNs were counted. The density of SGNs was calculated by dividing the SGN counts by the volume of the respective Rosenthal canals and expressed as SGN/ $10^5$ $\mu m^3$. Quantification of SGN density in D. Overall E. Base F. Middle, and G. Apical turn of the spiral ganglion. Error bars indicate SEM. Statistical analysis was performed using two-way ANOVA with Tukey’s multiple comparisons.
