## Supplementary figures and images for "Cochlear implants with dexamethasone-eluting electrode arrays reduce foreign body response in a murine model of cochlear implantation and human subjects"

### Supplementary Figure S1: Quality Control (QC) metrics for single cell RNA-Seq of CD11b+ immune cells.

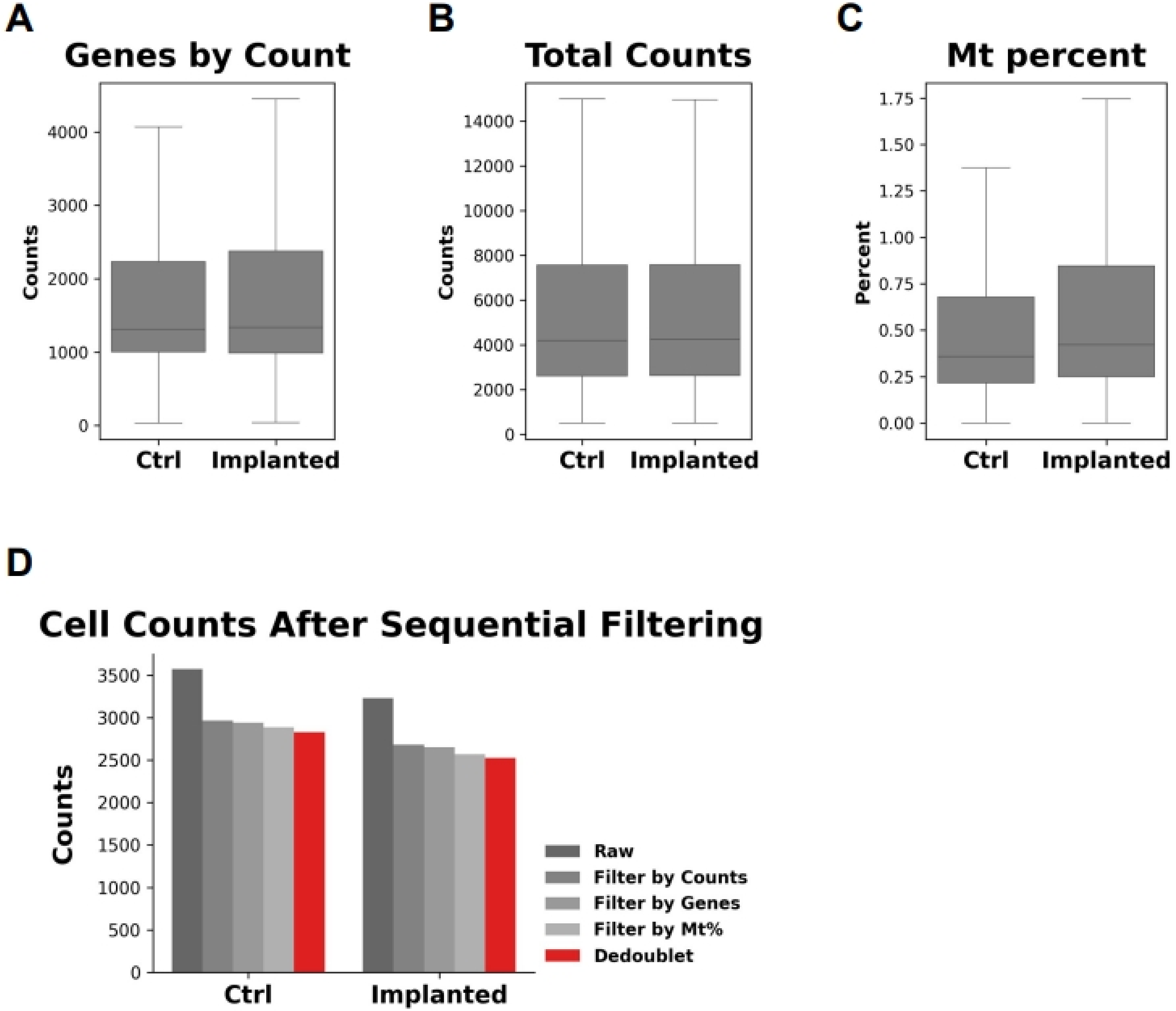

### Supplementary Figure S2: Quantification of macrophage infiltration following cochlear implantation in the middle, apical cochlear turns

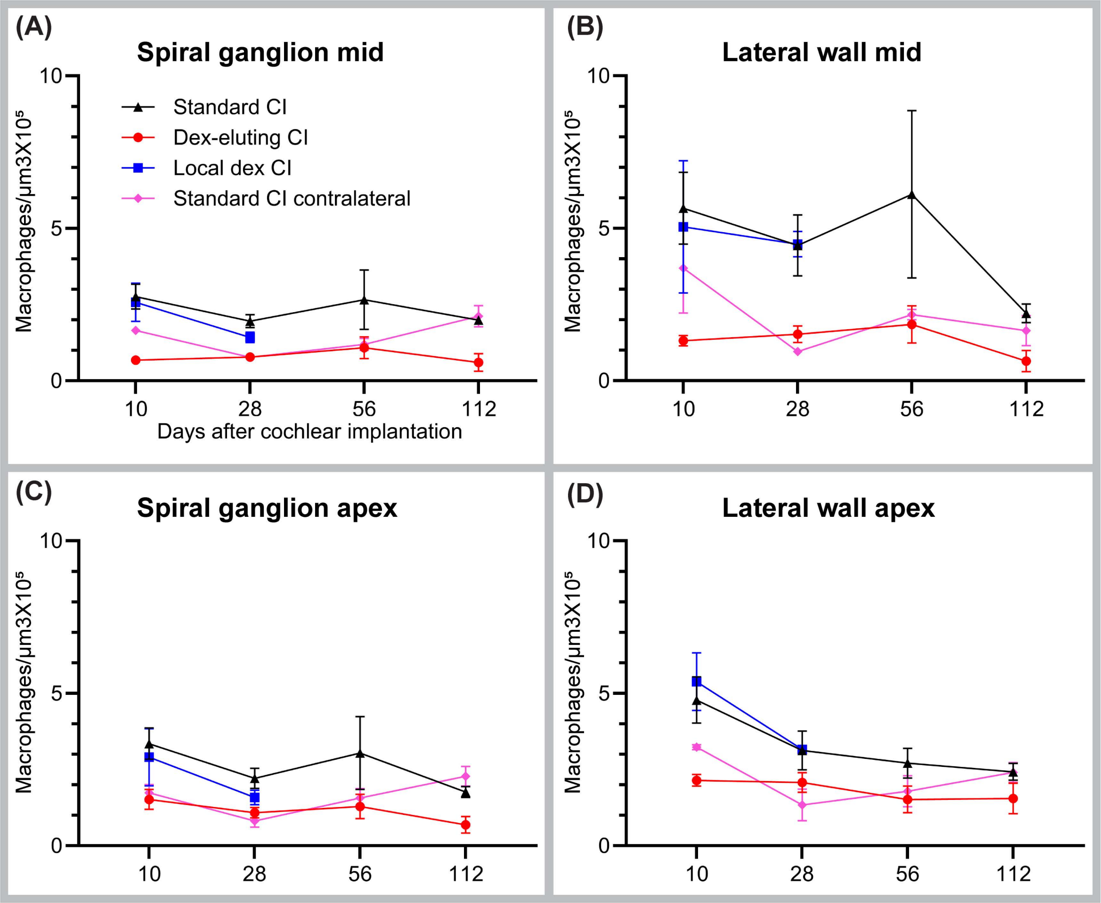

### Supplementary Figure S3: Quantification of cellular density in the spiral ganglia, lateral walls of Cochlea following implantation

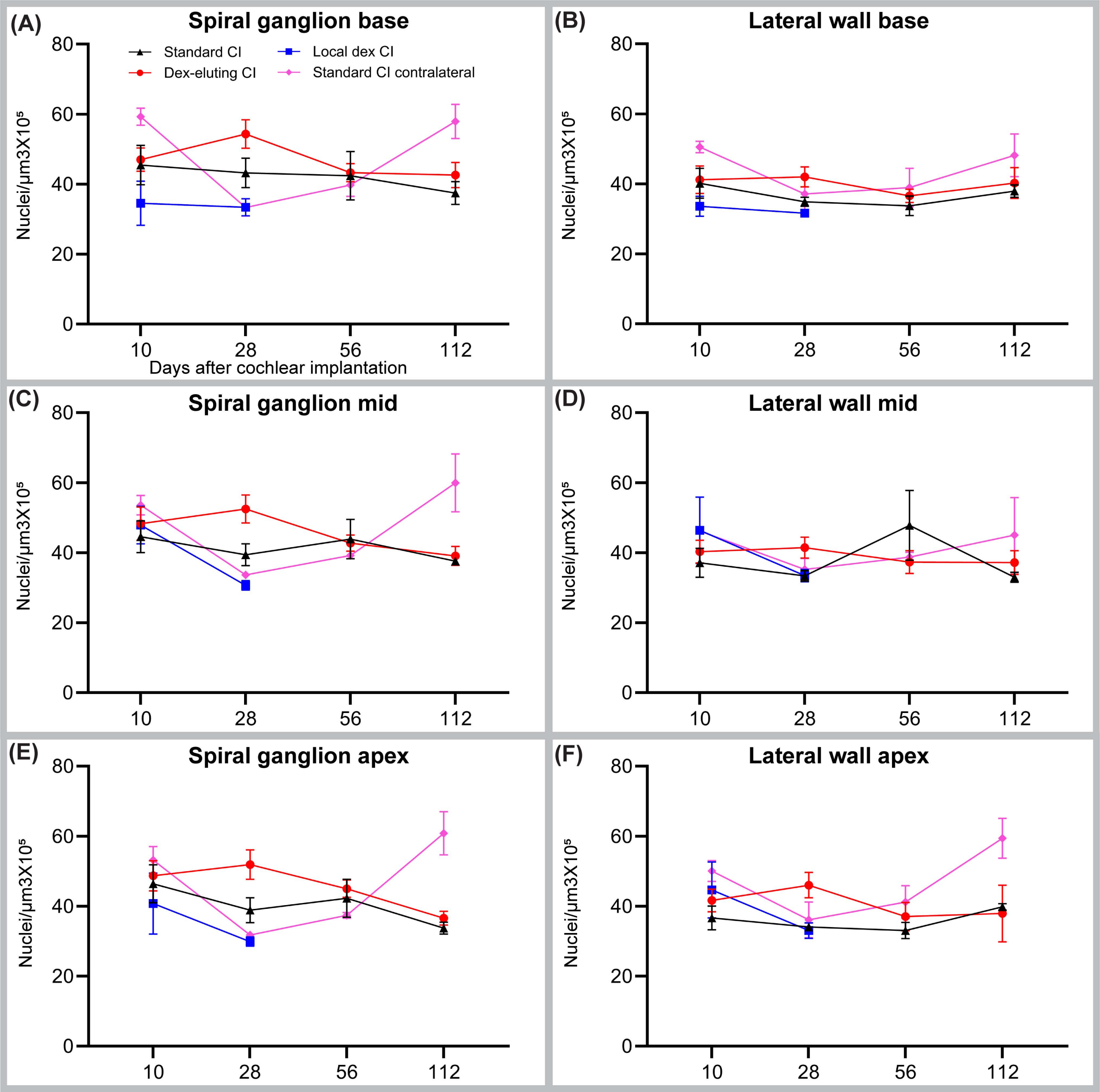

### Supplementary Figure S4: CX3CR1+ MHCII+ macrophages in implanted cochlea

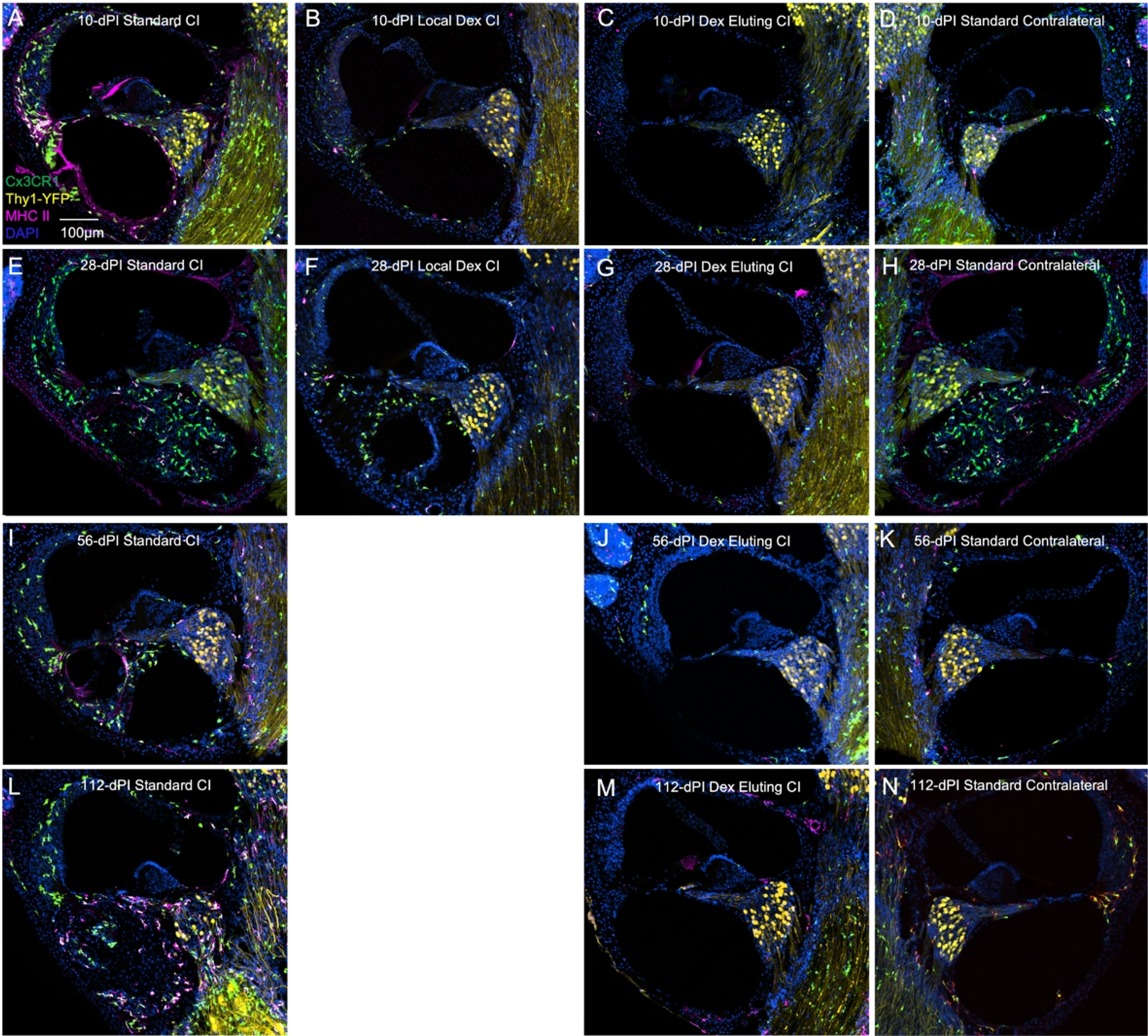

### Supplementary Figure S5: Quantification of CX3CR1+ MHCII+ macrophages in implanted cochlea

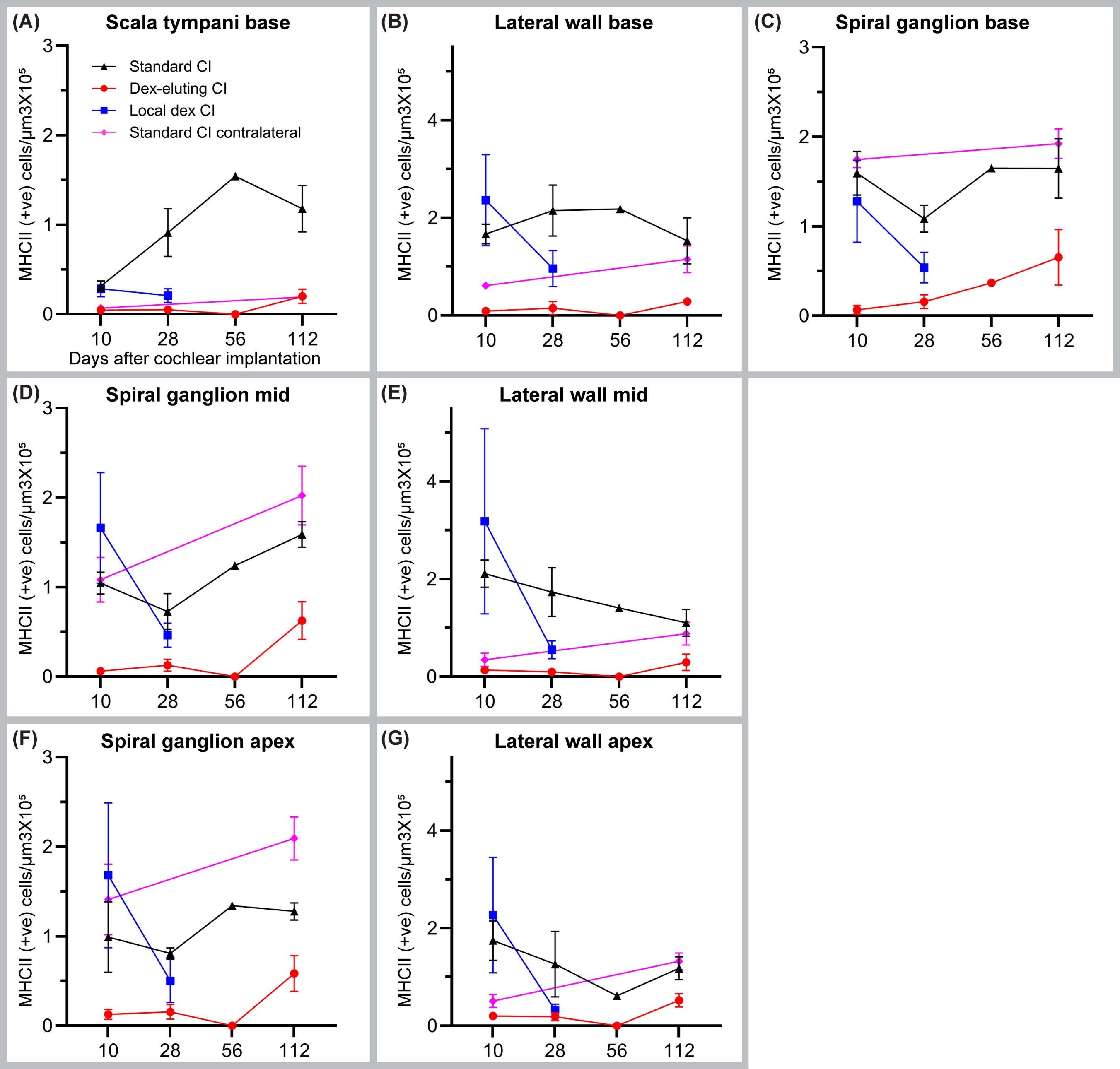

### Supplementary Figure S6: Spiral ganglion neurons (SGN) density following cochlear implantation

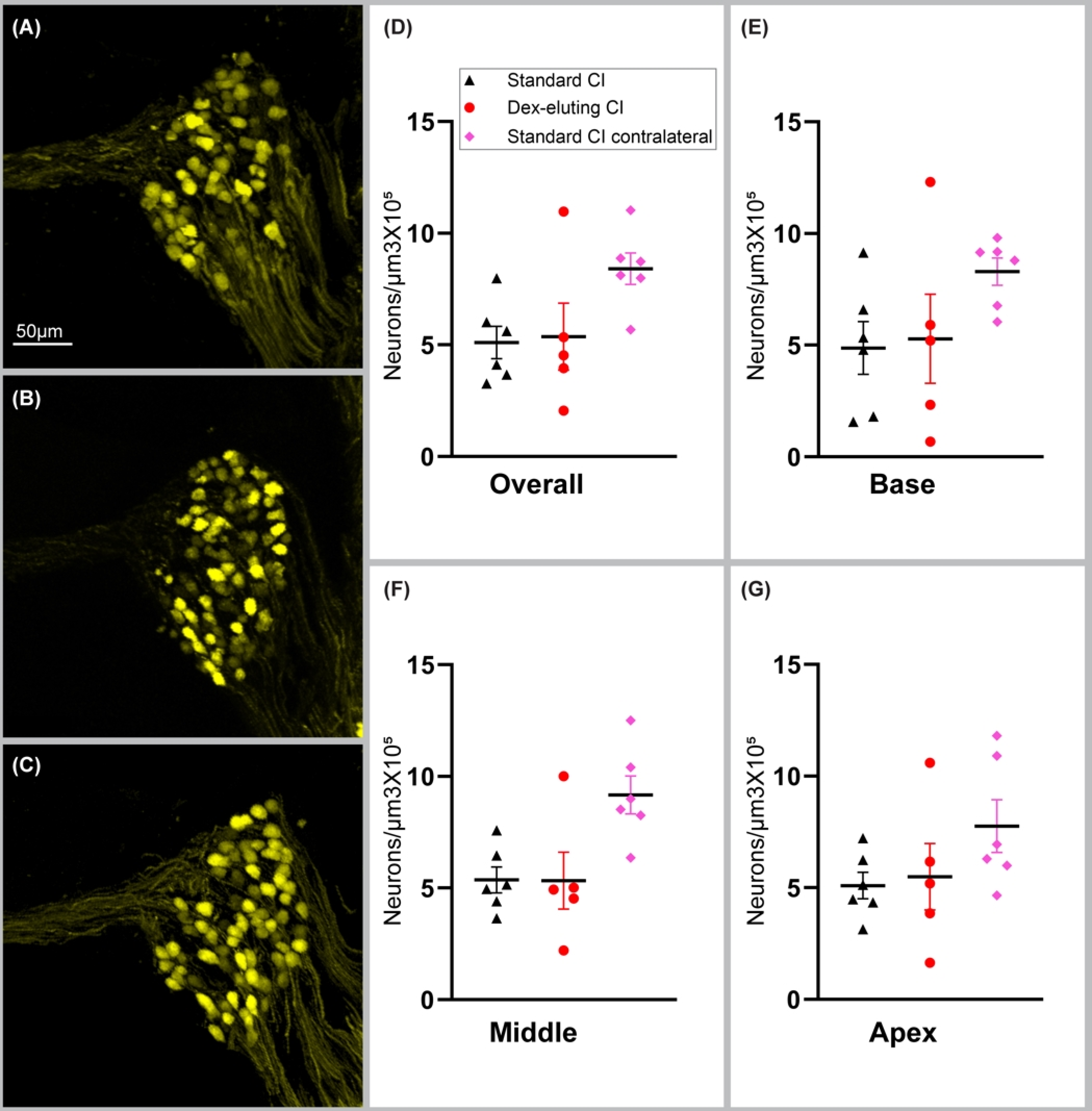
